# SARS-CoV-2 seroprevalence in Germany - a population based sequential study in five regions

**DOI:** 10.1101/2021.05.04.21256597

**Authors:** Daniela Gornyk, Manuela Harries, Stephan Glöckner, Monika Strengert, Tobias Kerrinnes, Gerhard Bojara, Stefanie Castell, Kerstin Frank, Knut Gubbe, Jana-Kristin Heise, Pilar Hernandez, Oliver Kappert, Winfried Kern, Thomas Illig, Norman Klopp, Henrike Maaß, Julia Ortmann, Barbora Kessel, Gottfried Roller, Monike Schlüter, Torsten Tonn, Michael Ziemons, Yvonne Kemmling, Berit Lange, Gérard Krause

## Abstract

Prevalence of SARS-CoV-2 antibodies is an essential indicator to guide measures. Few population-based estimates are available in Germany. We determine seroprevalence allowing comparison between regions, time points, socio-demographic and health-related factors.

MuSPAD is a sequential multi-local seroprevalence study. We randomly recruited adults in five counties with differing cumulative SARS-CoV-2 incidence July 2020 - February 2021. Serostatus was determined using Spike S1-specific IgG ELISA. We determined county-wise proportions of seropositivity. We assessed underestimation of infections, county and age specific infection fatality risks, and association of seropositivity with demographic, socioeconomic and health factors.

We found seroprevalence of 2.4 % (95%CI: 1.8-3.1%) for Reutlingen in June 2020 (stage 1) which increased to 2.9% (95%CI: 2.1-3.8%) in October (stage 2), Freiburg stage 1 1.5% (95% CI: 1.1-2.1%) vs. 2.5% (95%CI: 1.8-3.4%), Aachen stage 1 2.3% (95% CI: 1.7-3.1%) vs. 5.4% (95%CI: 4.4-6.6%), Osnabrück 1.3% (95% CI: 1.0-1.9%) and Magdeburg in Nov/Dec 2020. 2.4% (95%CI 1.9-3.1%). Number needed to quarantine to have one infected person quarantined was 8.2. The surveillance detection ratio (SDR) between number of infections based on our results and number reported to health authorities ranged from 2.5-4.5. Participants aged 80+ had lower SDR. Infection fatality estimates ranged from 0.2-2.4%. Lower education was associated with higher, smoking with lower seropositivity.

Seroprevalence remained low until December 2020 with high underdetection. The second wave from November 2020 to February 2021 resulted in additional 2-5% of the population being infected. Detected age specific differences of SDR should be taken into account in modelling and forecasting COVID-19 morbidity.

**Highlights:** *Evidence before this study:* Seroepidemiological surveys on SARS-CoV-2 are a useful tool to track the transmission during the epidemic. We searched PubMed/the pre-print server medRxiv and included web-based reports from German health organizations using the keywords “seroprevalence”, “SARS-CoV-2”, “Germany” and similar other English and German terms in the period from January 1st, 2020 until March 2021. We identified 30 published studies in Germany which mostly report low SARS-CoV-2 seroprevalence (<5%). Most of these surveys were so-called hotspot studies which assessed seroprevalence after localized outbreaks or examined seroprevalence of specific population groups such as e.g. medical staff. Few studies are either population-based or blood donor-based, but do not allow comparisons between regions. To date, we only consider the Corona sub-study of the Rhineland study similar to MuSPAD. It reports a low SARS-CoV-2 seroprevalence (46/4755; 0.97%; 95% CI: 0.72−1.30). Based on this, almost the entire German population remained susceptible to a SARS-CoV-2 infection by the end of 2020.

*Added value of this study:* We provide the first comprehensive, high-precision multi-region population-based SARS-CoV-2 seroprevalence study with representative sampling following the WHO protocol in Germany. By measuring SARS-CoV-2 IgG, we explore immunity at regional and national level over time. We also assess risk factors and sample each region twice, which permits to monitor seroprevalence progression throughout the epidemic in different exemplary German regions.

*Implications of all the available evidence:* Our results show low seroprevalence (<3%) until Mid-December 2020 in all regions. While estimates in Reutlingen, Aachen, Freiburg and Osnabrück reflect low seroprevalence mostly after the first wave, the survey in Magdeburg cumulatively already represents the beginning of the second wave. The number needed to quarantine to ensure one infected person was quarantined was 8.2 in our study. We also show that for the first wave reported infections reflected overall around 25% of those actually infected rising to 40-50% in the second wave. A slightly raised infection risk could be shown for persons with lower education.

## Introduction

More than one year into the pandemic, information about the actual extent of SARS-CoV-2 infections in the German population is still largely based on the number of COVID-19 cases reported to local health care authorities based on regulations of the German Infection Protection Act (IfSG). In January 2020, the first case in Bavaria, Germany was detected ^1^. Shortly after, more cases occurred, many of which in North-Rhine Westphalia, linked to a carnival session ^2^. In the following months several clusters occurred in Lower Saxony in meat processing facilities^3^. The first epidemic wave in spring 2020 was characterized by comparably low age-standardized case fatality estimates^4^ and low excess mortality ^5^. In the beginning of the pandemic the infection management focused on outbreak investigations, hotspots and PCR screenings for specific populations like health care workers.

Consequently, current estimates of SARS-CoV-2 spread are not reliable as they do not include frequently occurring non-notified asymptomatic or mild SARS-COV-2 infections ^6 7^. In contrast, population-based cohort studies measuring IgG antibodies can shed light on the number of persons with prior SARS-CoV-2 exposure independent of clinical manifestation and determine age specific infection fatality risk. Precise knowledge and quantification of SARS-CoV-2 seroprevalence and its development over time can aid to judge the effectiveness of population-based interventions, direct future preventive strategies and guide vaccination strategies to build herd immunity independent of natural exposure.

To date there are over 30 published seroepidemiological studies in Germany, with only one general focussing on the population not based in hotspots ^8^, two with populations consisting of children^9 10^ and five studies among blood donors ^11 12^. One of the largest studies “CORONA-MONITORING local”^13^ conducted by the Robert Koch Institute, focused on recent hotspots. In Kupferzell the seroprevalence was 7.7% which implies 3.9 times more past infections than cases reported by the health authorities. Similarly, in Bad Feilnbach the seroprevalence equals 2.6 times more infections than previously known. So far, the above mentioned existing sub-cohort study conducted in Bonn, targeted the general population without aforementioned focus ^8^.

The sample size and methodology of these studies do not allow to provide population group specific estimates from larger non-hotspot counties and regions within Germany. To address the lack of data on the extent of SARS-CoV-2 dissemination in the general population and to allow for a better comparison with other European countries despite differing surveillance systems, we established MuSPAD (Multilocal and Serial Prevalence Study of Antibodies against SARS-2 Coronavirus in Germany).

## Methods

### Study design, participants and setting

We followed the WHO protocol^14^ for SARS-CoV-2 seroprevalence studies and STROBE as a reporting guideline for observational studies ^15^. MuSPAD is an ongoing population-based seroepidemiological observational study consisting of successive cross-sectional studies with longitudinal components (follow ups within three months). Each follow-up comprises a new cross-sectional study. Main data collection period is from July 2020 until August 2021.

Residents’ registration offices drew a random sample taking into account age (≥18 years), gender and spatial distributions ^16^.

Individuals who received a written invitation, provided written informed consent and did not have contraindications for giving a blood sample were eligible for the study. All study materials were provided in German. Individuals with suspected or confirmed SARS-CoV-2 infections were not excluded.

The spatial and temporal patterns of COVID 19 incidence were quite different within Germany. Initially heavily impacted was the south of Germany which is why we chose the urban and rural area of Freiburg including Breisgau-Hochschwarzwald and a county called Reutlingen in the south west. These belong to the state of Baden-Württemberg and had reported early high notified case incidences. Cumulative incidence approached 500/100,000 population in June 2020.

Freiburg is a city of about 250,000 inhabitants with a surrounding rural area of Breisgau-Hochschwarzwald (hereafter referred to as Freiburg). Its defining feature in the epidemic in March 2020 was a geographic closeness to one of the hardest hit regions in France, the Alsace. It was the first city in Germany to instate severe contact restrictions, already on 18^th^ of March 2020. Reutlingen is a city of around 115,000 inhabitants in the south-west of Germany and has a surrounding rural area of Swabian Alb. The industrialised county of Reutlingen overall has a population of around 280,000 residents.

The first COIVD-19 wave in Germany happened between March and the beginning of April 2020, in particular in north-west hotspots attributed to the carnival season. The StädteRegionAachen (around 560.000 inhabitants) (hereafter referred to as Aachen) was chosen because of its strong carnival tradition. It consists of one larger city, several rural regions, and mid-level cities and includes border regions to Belgium. When sampling a second time in Aachen, some of the residents already had been vaccinated. In the further course several COVID-19 clusters occurred in Lower Saxony, Northern Germany. We selected the urbanized and rural area of Osnabrück to sample the general population. The rural region of Osnabrück is home to several meat producing industries, where outbreaks of SARS-CoV-2 were registered from March to July 2020 (^3^).

In the eastern part of Germany case numbers increased only slowly. Here, we chose Magdeburg with its 236.000 inhabitants as the capital of the state of Saxony-Anhalt which had only scattered outbreaks during June 2020 but started to have a second wave of cases in November 2020 (Figure 1).

**Figure 1:**
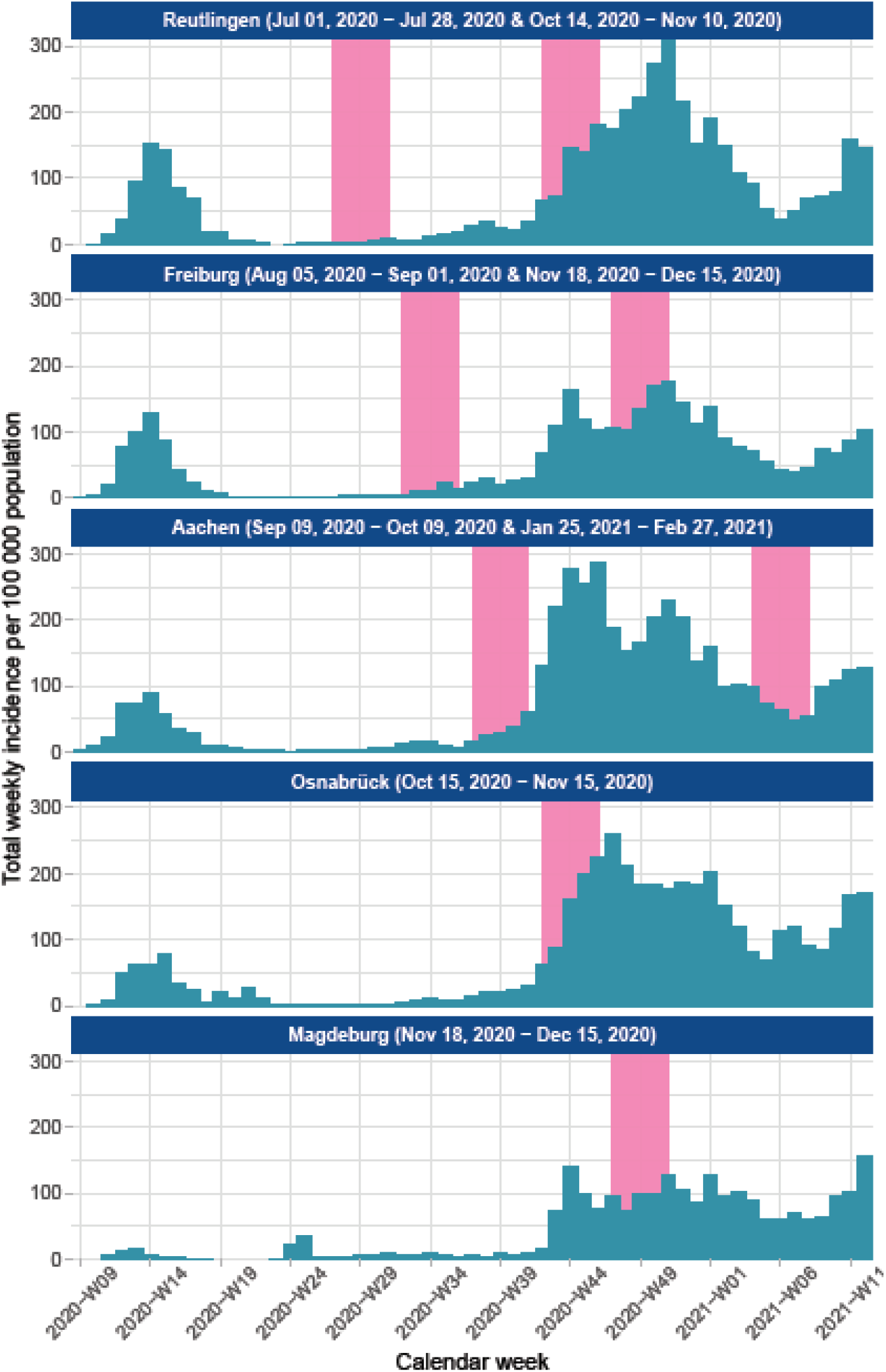
MuSPAD sampling time per site compared with reported cases Source @RKI in Germany, July 2020-February 2021, Data for rural area and city Osnabrück and Freiburg aggregated.

In each location a study centre was set up to enable questionnaire based interviews and blood collection of around 100-150 individuals per day. Study centres followed Standardized Operating Procedures (SOPs). Invited persons booked an appointment either in the study centre, or – if symptomatic, immobile or frail - with a mobile study team. Informed consent was given after explanation of study procedures and risks by trained study staff with a medical doctor present for questions. Subsequently, a questionnaire was administered and a blood sample was taken. The study participant was made aware of a follow-up questionnaire available on paper or online (web or mobile app).

For data collection and follow-up the application PIA (**P**rospective Monitoring and Management – **A**pp)^17^ was used. PIA enables the consolidation of data from different sources (study staff, participants) in real-time, simplifying data management and improving data quality. PIA complies with German data protection and IT security requirements and has been regulary evaluated by penetration tests.

Additional information on recruitment procedures, questionnaires, laboratory procedures, and data protection are shared in the Supplement section.

### Sample preparation and lab analysis

A venous blood sample of 9 mL using barcoded serum-gel monovettes was performed at the test centre or at home. After centrifugation (2500g, 10min at ambient temperature) to separate serum from cellular blood components, samples were stored at 4-8°C until analysis at the Institute for Transfusion medicine in Plauen. Spike S1 -specific IgGs were measured using the semi-quantitative SARS-CoV-2 IgG ELISA from Euroimmun (Lübeck, Germany) according to the manufactures instruction. The remaining serum was aliquoted, frozen and stored at the Hannover Unified Biobank (HUB).

### Data analysis and data analysis plan

We estimated prevalence ratios using a 95% confidence interval (CI) using crude and population weighted^18^ subgroups for age and sex per county. We further analysed the influence of other factors - such as comorbidities, residential area and work related factors - in multivariable logistic regression analysis with variables determined in advance accounting for clustering by study site. Expected infections were calculated for each study site as the product of seroprevalence per site multiplied by inhabitants. We computed a ratio for the sensitivity of the surveillance system to detect infections (Surveillance Detection Ratio), taking the number of persons of a study region having ever been notified as SARS-CoV-2 cases (from onset of the pandemic until 14 days before blood samples were taken), and dividing it by the number of persons expected to be infected according to seroprevalence estimates as calculated above. We calculated the number needed to quarantine to find one further infection as 1 (Prevalence in the population quarantined – Prevalence in the population not quarantined). Source data for deaths were daily death counts provided by RKI with a cut off at the start of the study (14 days after cases were counted). We compared this infection fatality risk calculated as notified deaths at the start of the study over expected number of persons infected according to seroprevalence to each study site. Analyses were done using STATA (Version 14 and 16) and R Version 4.0.2.

We also provide estimates of seropositivity and their uncertainties accounted for the test performance by deriving their values from Bayesian hierarchical models (as in^19^). Such models both reflect the influence of test’s sensitivity and specificity on the observed numbers of seropositive study participants and account for the uncertainties tied to these test’s characteristics determined from the diagnostic accuracy. These estimates were provided as a sensitivity analysis and not used for the main estimations (Supplement).

### Ethics

MuSPAD complies with all relevant laws and declarations (EU Charter of Fundamental Rights, Biomedical Convention of the Council of Europe and additional protocols, the CIOMS guidelines and the Helsinki Declaration), ethical approval was given on 21.06.2020 by the Ethics Committee of the Hannover Medical School (Ethics approval no 9086_BO_S_2020).

## Results

### Characteristics of study participants

We recruited 13,045 participants for stage 1 and 6,160 new participants (excluding follow-up participants) in stage 2 (Figure 2) with age ranging from 18 to 99 years (median 51), 55% of which female. Proportions of self-reported chronic conditions were similar across regions and stages (overall 26.0% hypertension, 10.4% cardiovascular disease, 8.3% chronic lung disease, 6.1% diabetes and 2.5% cancer (stage 1)) except for Magdeburg with slightly higher proportions for diabetes, hypertension and cardiovascular disease (Tabel 2). Proportion of daily smokers ranged from 10.1% in Freiburg to 16.8% in Reutlingen. Seventy-five percent of the participants shared a household with other adults, 18.4% lived alone and 24.4% shared their household with children. The proportion of individuals with higher education ranged from 45.2% in Osnabrück to 70.2% in Freiburg. Fifty-nine percent reported having experienced changes in their work during the pandemic. For stage 2 (Reutlingen 2, Freiburg 2 and Aachen 2) the demographic characteristics (female 54.7%, mean age 50.9) were similar to stage 1. There were no noteworthy differences in chronic conditions (23.4% hypertension, 8.9% cardiovascular disease, 8.1% chronic lung disease, 5.0% diabetes and 2.7% cancer) or daily smoking (RT2: 13.9%, FR2: 10% and AC2: 13%) between stages.

**Figure 2:**
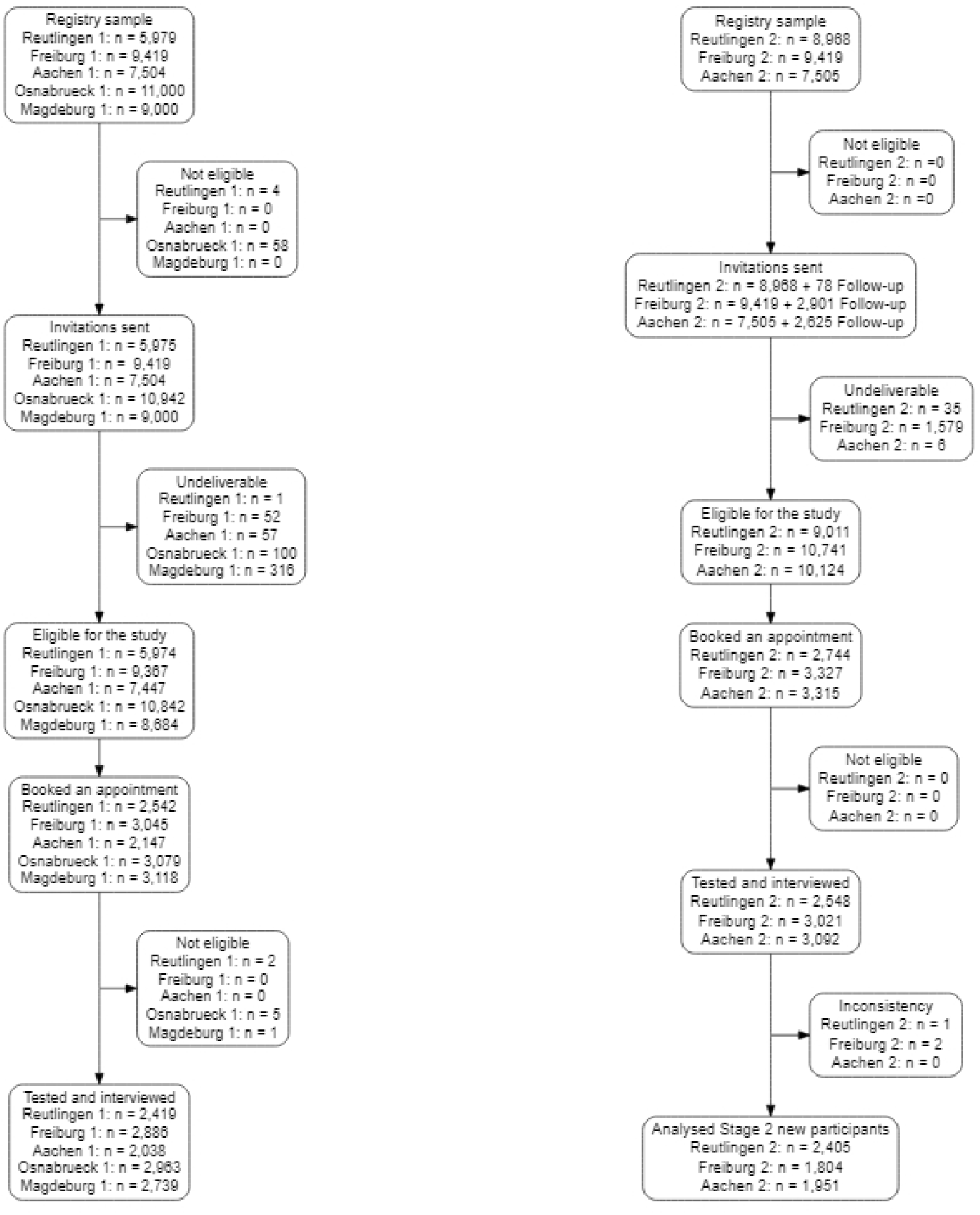
STROBE flow chart of MuSPAD participants in five study regions for stage 1 and three study regions for stage 2 excluding follow up, Germany July 2020-February 2021

**Table 1:** Comparison of participants’ characteristics from different study sites (Reutlingen (RT), Freiburg (FR), Aachen (AC) for the first phase, July-December 2020

| Characteristic | Participants<br>RT<br>N (%) | Participants<br>FR<br>N (%) | Participants<br>AC<br>N (%) | Participants<br>OS<br>N (%) | Participants<br>MD<br>N (%) | Total<br>N (%) |
| --- | --- | --- | --- | --- | --- | --- |
| Population | 2419 | 2886 | 2038 | 2963 | 2739 | 13045 |
| **Area** |  |  |  |  |  |  |
| Urban | 1938 (80.1%) | 2434 (84.3%) | 1246 (61.1%) | 707 (23.9%) | 2532 (92.4%) | 8857 (67.9%) |
| Rural | 481 (19.9%) | 452 (15.7%) | 792 (38.9%) | 2256 (76.1%) | 207 (7.6%) | 4188 (32.1%) |
| Age (Mean, SD) | 50.6 (17.4) | 48.6 (17.6) | 50.4 (17.6) | 51.2 (16.2) | 53.7 (17.8) | 50.9 (17.4) |
| **Age group, years** |  |  |  |  |  |  |
| 18-25 | 242 (10.4%) | 264 (9.3%) | 202 (10.0%) | 229 (7.8%) | 144 (5.3%) | 1081 (8.4%) |
| 26-45 | 614 (26.4%) | 1018 (36.0%) | 582 (28.7%) | 784 (26.8%) | 803 (29.7%) | 3801 (29.7%) |
| 46-65 | 1004 (43.2%) | 998 (35.3%) | 800 (39.5%) | 1345 (45.9%) | 974 (36.0%) | 5121 (40.0%) |
| 66-79 | 346 (14.9%) | 427 (15.1%) | 345 (17.0%) | 463 (15.8%) | 570 (21.1%) | 2151 (16.8%) |
| > 79 | 118 (5.1%) | 123 (4.3%) | 96 (4.7%) | 107 (3.7%) | 211 (7.8%) | 655 (5.1%) |
| Missing | 95 (3.9%) | 56 (2.3%) | 13 (0.5%) | 35 (1.5%) | 37 (1.5%) | 236 (9.8%) |
| **Sex** |  |  |  |  |  |  |
| Male | 1075 (44.9%) | 1226 (42.8%) | 912 (44.9%) | 1321 (45.1%) | 1267 (46.5%) | 5801 (44.8%) |
| Female | 1319 (55.1%) | 1638 (57.2%) | 1120 (55.1%) | 1611 (54.9%) | 1455 (53.5%) | 7143 (55.2%) |
| Missing | 25 (1.03%) | 22 (0.76%) | 6 (1.05%) | 31 (0.62%) | 17 (0.62%) | 101 (0.77%) |
| **Predisposing conditions** |  |  |  |  |  |  |
| Diabetes | 147 (6.2%) | 115 (4.0%) | 103 (5.1%) | 152 (5.1%) | 275 (10.1%) | 792 (6.1%) |
| Hypertension | 520 (22.1%) | 524 (18.2%) | 503 (24.7%) | 792 (26.8%) | 1032 (37.8%) | 3371 (26.0%) |
| Cardiovascular condition | 188 (8.0%) | 242 (8.4%) | 243 (11.9%) | 295 (10.0%) | 386 (14.1%) | 1354 (10.4%) |
| Chronic lung disease | 155 (6.6%) | 243 (8.5%) | 198 (9.7%) | 242 (8.2%) | 243 (8.9%) | 1081 (8.3%) |
| Cancer | 52 (2.2%) | 67 (2.3%) | 40 (2.0%) | 67 (2.3%) | 95 (3.5%) | 321 (2.5%) |
| Immunosuppressed | 61 (2.6%) | 106 (3.7%) | 85 (4.2%) | 95 (3.2%) | 110 (4.0%) | 457 (3.5%) |
| Missing | 63 (2.6%) | 11 (0.38%) | 0 | 7 (0.2%) | 6 (0.24%) | 87 (0.67%) |
| **Tobacco consumption** |  |  |  |  |  |  |
| Never smoker | 1258 (53.3%) | 1448 (50.4%) | 1028 (50.5%) | 1402 (47.4%) | 1288 (47.1%) | 6424 (49.6%) |
| Previous smoker | 592 (25.1%) | 935 (32.6%) | 639 (31.4%) | 963 (32.6%) | 879 (32.1%) | 4008 (30.9%) |
| Occasional smoker | 113 (4.8%) | 198 (6.9%) | 104 (5.1%) | 144 (4.9%) | 134 (4.9%) | 693 (5.3%) |
| Regular smoker | 396 (16.8%) | 291 (10.1%) | 264 (13.0%) | 446 (15.1%) | 436 (15.9%) | 1833 (14.1%) |
| Missing | 60 (2.5%) | 14 (0.5%) | 3 (0.1%) | 8 (0.3%) | 2 (0.1%) | 87 (0.7%) |
| **Living conditions** |  |  |  |  |  |  |
| Flat/house | 2326 (98.8%) | 2830 (98.5%) | 2021 (99.2%) | 2941 (99.7%) | 2695 (99.4%) | 12813 (99.1%) |
| Assisted/other | 28 (1.2%) | 42 (1.5%) | 17 (0.8%) | 9 (0.3%) | 15 (0.6%) | 111 (0.9%) |
| **Household size, residents** |  |  |  |  |  |  |
| Lives alone | 353 (15.1%) | 561 (19.5%) | 420 (20.6%) | 430 (14.5%) | 622 (22.7%) | 2386 (18.4%) |
| Lives with 1-3 persons | 1831 (78.2%) | 2097 (73.0%) | 1450 (71.2%) | 2306 (78.0%) | 2028 (74.1%) | 9712 (75.0%) |
| Lives with > 3 persons | 156 (6.7%) | 215 (7.5%) | 167 (8.2%) | 221 (7.5%) | 88 (3.2%) | 847 (6.5%) |
| Missing | 79 (3.3%) | 13 (0.5%) | 1 (0.0%) | 6 (0.2%) | 1 (0.0%) | 100 (0.8%) |
| **Children in the household** |  |  |  |  |  |  |
| None | 1743 (74.5%) | 2166 (75.4%) | 1541 (78.6%) | 2110 (71.4%) | 2159 (78.9%) | 9719 (75.5%) |
| Yes, youngest 0-10 years | 345 (14.7%) | 462 (16.1%) | 275 (14.0%) | 458 (15.5%) | 408 (14.9%) | 1948 (15.1%) |
| Yes, youngest 11-18 years | 252 (10.8%) | 245 (8.5%) | 145 (7.4%) | 389 (13.2%) | 171 (6.2%) | 1202 (9.3%) |
| Missing | 79 (3.3%) | 13 (0.5%) | 77 (3.8%) | 6 (0.2%) | 1 (0.0%) | 176 (1.3%) |
| **Lives with** |  |  |  |  |  |  |
| Living alone | 353 (15.1%) | 561 (19.5%) | 420 (20.6%) | 430 (14.5%) | 622 (22.7%) | 2386 (18.4%) |
| Lives with adults | 1387 (59.3%) | 1601 (55.7%) | 1168 (57.3%) | 1674 (56.6%) | 1537 (56.1%) | 7367 (56.9%) |
| Lives with kids youngest <5 | 219 (9.4%) | 297 (10.3%) | 165 (8.1%) | 292 (9.9%) | 204 (7.5%) | 1177 (9.1%) |
| Lives with kids youngest >=5 | 381 (16.3%) | 414 (14.4%) | 284 (13.9%) | 561 (19.0%) | 375 (13.7%) | 2015 (15.6%) |
| **Education** |  |  |  |  |  |  |
| Certificate after 9 years | 478 (19.8%) | 277 (9.6%) | 294 (14.4%) | 561 (18.9%) | 248 (9.1%) | 1858 (14.2%) |
| Certificate after 10 years | 662 (27.4%) | 552 (19.1%) | 374 (18.4%) | 1041 (35.1%) | 1018 (37.2%) | 3647 (28.0%) |
| Higher education | 1170 (48.4%) | 2026 (70.2%) | 1348 (66.1%) | 1338 (45.2%) | 1446 (52.8%) | 7328 (56.2%) |
| None | 36 (1.5%) | 9 (0.3%) | 16 (0.8%) | 8 (0.3%) | 11 (0.4%) | 80 (0.6%) |
| Missing | 73 (3.0%) | 22 (0.8%) | 6 (0.3%) | 15 (0.5%) | 16 (0.6%) | 132 (1.0%) |
| **Occupation** |  |  |  |  |  |  |
| Employed | 1354 (57.9%) | 1706 (59.5%) | 1128 (55.9%) | 1847 (62.7%) | 1485 (54.7%) | 7520 (58.4%) |
| Self employed | 132 (5.6%) | 233 (8.1%) | 135 (6.7%) | 178 (6.0%) | 134 (4.9%) | 812 (6.3%) |
| Unemployed | 41 (1.8%) | 36 (1.3%) | 47 (2.3%) | 34 (1.2%) | 61 (2.2%) | 219 (1.7%) |
| Retraining / Voluntary | 9 (0.4%) | 7 (0.2%) | 4 (0.2%) | 5 (0.2%) | 7 (0.3%) | 32 (0.2%) |
| Apprenticeship | 45 (1.9%) | 30 (1.0%) | 13 (0.6%) | 10 (0.3%) | 14 (0.5%) | 112 (0.9%) |
| Parental / other leave | 75 (3.2%) | 74 (2.6%) | 54 (2.7%) | 53 (1.8%) | 32 (1.2%) | 288 (2.2%) |
| Pupil / Retired | 682 (29.2%) | 780 (27.2%) | 637 (31.6%) | 821 (27.8%) | 984 (36.2%) | 3904 (30.3%) |
| Missing | 81 (3.3%) | 20 (0.7%) | 20 (1.0%) | 15 (0.5%) | 22 (0.8%) | 158 (1.2%) |
| **Changes due to COVID** |  |  |  |  |  |  |
| No changes | 1443 (59.6%) | 1506 (52.2%) | 1160 (56.2%) | 1772 (59.8%) | 1828 (66.7%) | 7709 (59.1%) |
| Work from home | 461 (20.1%) | 520 (17.8%) | 603 (29.5%) | 623 (21%) | 427 (20.6%) | 2634 (21.4%) |
| Reduction work | 454 (18.8%) | 472 (16.4%) | 245 (12.0%) | 390 (13.2%) | 193 (7.0%) | 1754 (13.4%) |
| Increase work | 192 (7.9%) | 315 (10.9%) | 206 (10.1%) | 291 (9.8%) | 230 (8.4%) | 1234 (9.5%) |
| Short time work | 364 (37.0%) | 249 (17.3%) | 127 (13.2%) | 356 (27.4%) | 175 (17.0%) | 1271 (22.2%) |
| Termination | 11 (1.1%) | 33 (2.3%) | 15 (1.6%) | 25 (1.9%) | 15 (1.5%) | 99 (1.7%) |
| New workplace | 3 (0.3%) | 51 (3.5%) | 21 (2.2%) | 44 (3.4%) | 41 (4.0%) | 160 (2.8%) |
| Other consequences | 0 (0.0%) | 88 (6.1%) | 108 (11.2%) | 130 (10.0%) | 134 (13.0%) | 460 (8.0%) |

**Table 2:**
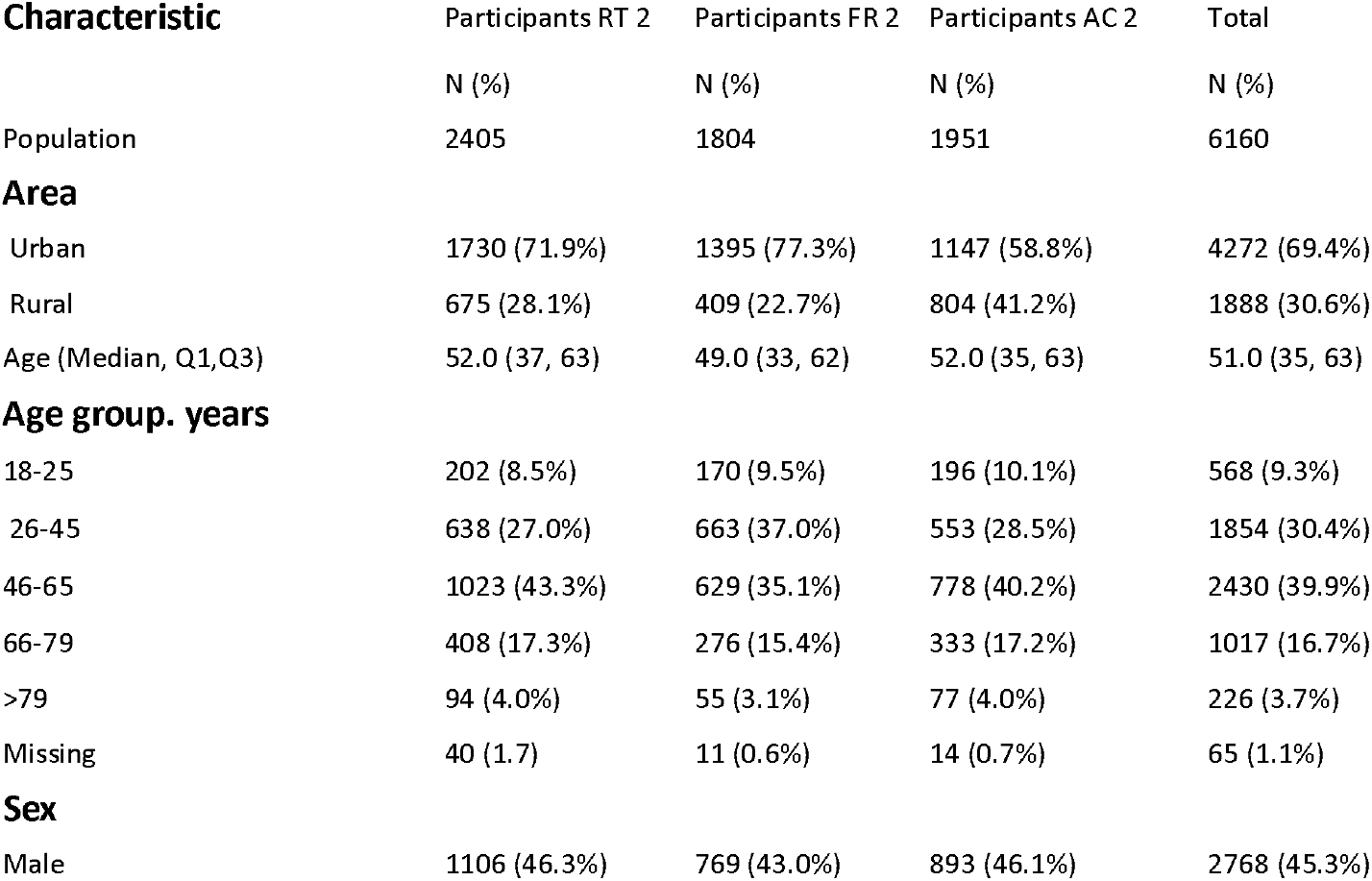
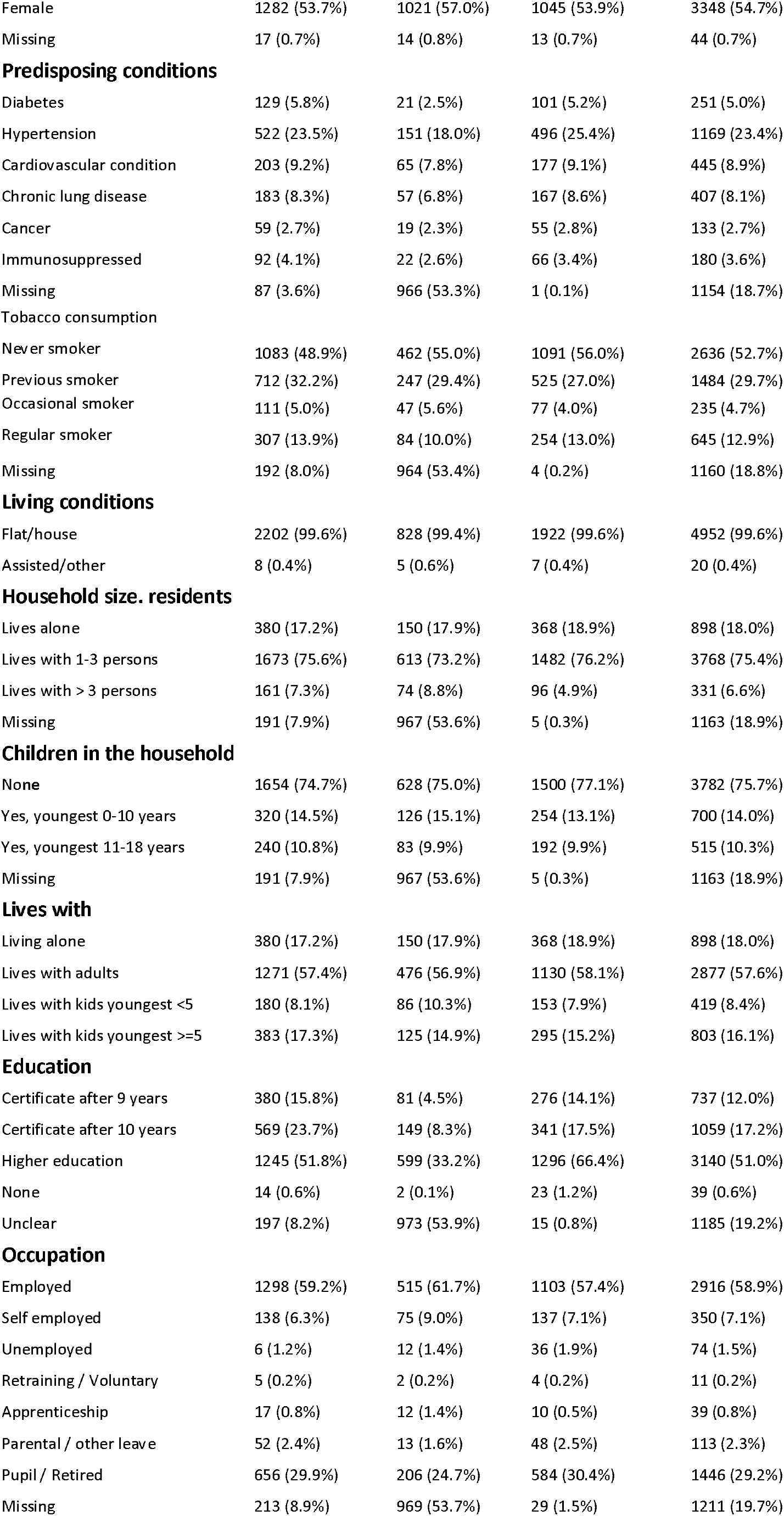
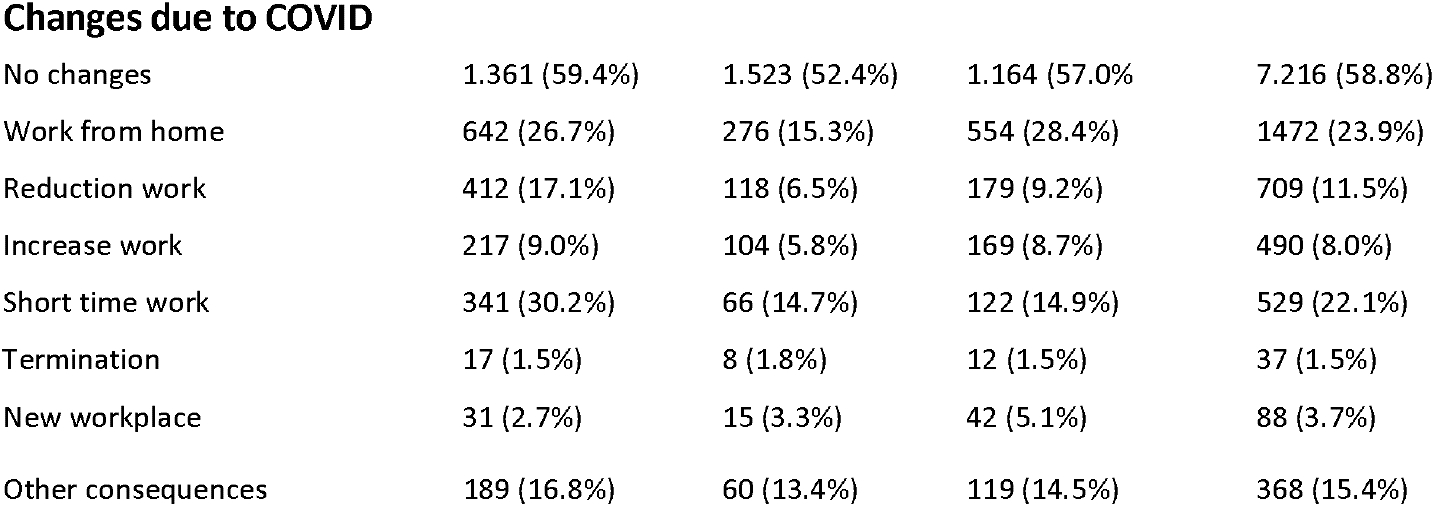
Comparison of sampled participants’ characteristics in different study population (Reutlingen (RT), Freiburg (FR), Aachen (AC) for the second phase, October-February 2020-2021

| Characteristic | Participants RT 2 | Participants FR 2 | Participants AC 2 | Total |
| --- | --- | --- | --- | --- |
|  | N (%) | N (%) | N (%) | N (%) |
| Population | 2405 | 1804 | 1951 | 6160 |
| **Area** |  |  |  |  |
| Urban | 1730 (71.9%) | 1395 (77.3%) | 1147 (58.8%) | 4272 (69.4%) |
| Rural | 675 (28.1%) | 409 (22.7%) | 804 (41.2%) | 1888 (30.6%) |
| Age (Median, Q1,Q3) | 52.0 (37, 63) | 49.0 (33, 62) | 52.0 (35, 63) | 51.0 (35, 63) |
| **Age group. years** |  |  |  |  |
| 18-25 | 202 (8.5%) | 170 (9.5%) | 196 (10.1%) | 568 (9.3%) |
| 26-45 | 638 (27.0%) | 663 (37.0%) | 553 (28.5%) | 1854 (30.4%) |
| 46-65 | 1023 (43.3%) | 629 (35.1%) | 778 (40.2%) | 2430 (39.9%) |
| 66-79 | 408 (17.3%) | 276 (15.4%) | 333 (17.2%) | 1017 (16.7%) |
| >79 | 94 (4.0%) | 55 (3.1%) | 77 (4.0%) | 226 (3.7%) |
| Missing | 40 (1.7) | 11 (0.6%) | 14 (0.7%) | 65 (1.1%) |
| **Sex** |  |  |  |  |
| Male | 1106 (46.3%) | 769 (43.0%) | 893 (46.1%) | 2768 (45.3%) |
| Female | 1282 (53.7%) | 1021 (57.0%) | 1045 (53.9%) | 3348 (54.7%) |
| Missing | 17 (0.7%) | 14 (0.8%) | 13 (0.7%) | 44 (0.7%) |
| **Predisposing conditions** |  |  |  |  |
| Diabetes | 129 (5.8%) | 21 (2.5%) | 101 (5.2%) | 251 (5.0%) |
| Hypertension | 522 (23.5%) | 151 (18.0%) | 496 (25.4%) | 1169 (23.4%) |
| Cardiovascular condition | 203 (9.2%) | 65 (7.8%) | 177 (9.1%) | 445 (8.9%) |
| Chronic lung disease | 183 (8.3%) | 57 (6.8%) | 167 (8.6%) | 407 (8.1%) |
| Cancer | 59 (2.7%) | 19 (2.3%) | 55 (2.8%) | 133 (2.7%) |
| Immunosuppressed | 92 (4.1%) | 22 (2.6%) | 66 (3.4%) | 180 (3.6%) |
| Missing | 87 (3.6%) | 966 (53.3%) | 1 (0.1%) | 1154 (18.7%) |
| **Tobacco consumption** |  |  |  |  |
| Never smoker | 1083 (48.9%) | 462 (55.0%) | 1091 (56.0%) | 2636 (52.7%) |
| Previous smoker | 712 (32.2%) | 247 (29.4%) | 525 (27.0%) | 1484 (29.7%) |
| Occasional smoker | 111 (5.0%) | 47 (5.6%) | 77 (4.0%) | 235 (4.7%) |
| Regular smoker | 307 (13.9%) | 84 (10.0%) | 254 (13.0%) | 645 (12.9%) |
| Missing | 192 (8.0%) | 964 (53.4%) | 4 (0.2%) | 1160 (18.8%) |
| **Living conditions** |  |  |  |  |
| Flat/house | 2202 (99.6%) | 828 (99.4%) | 1922 (99.6%) | 4952 (99.6%) |
| Assisted/other | 8 (0.4%) | 5 (0.6%) | 7 (0.4%) | 20 (0.4%) |
| **Household size. residents** |  |  |  |  |
| Lives alone | 380 (17.2%) | 150 (17.9%) | 368 (18.9%) | 898 (18.0%) |
| Lives with 1-3 persons | 1673 (75.6%) | 613 (73.2%) | 1482 (76.2%) | 3768 (75.4%) |
| Lives with > 3 persons | 161 (7.3%) | 74 (8.8%) | 96 (4.9%) | 331 (6.6%) |
| Missing | 191 (7.9%) | 967 (53.6%) | 5 (0.3%) | 1163 (18.9%) |
| **Children in the household** |  |  |  |  |
| None | 1654 (74.7%) | 628 (75.0%) | 1500 (77.1%) | 3782 (75.7%) |
| Yes, youngest 0- 10 years | 320 (14.5%) | 126 (15.1%) | 254 (13.1%) | 700 (14.0%) |
| Yes, youngest 11-18 years | 240 (10.8%) | 83 (9.9%) | 192 (9.9%) | 515 (10.3%) |
| Missing | 191 (7.9%) | 967 (53.6%) | 5 (0.3%) | 1163 (18.9%) |
| **Lives with** |  |  |  |  |
| Living alone | 380 (17.2%) | 150 (17.9%) | 368 (18.9%) | 898 (18.0%) |
| Lives with adults | 1271 (57.4%) | 476 (56.9%) | 1130 (58.1%) | 2877 (57.6%) |
| Lives with kids youngest <5 | 180 (8.1%) | 86 (10.3%) | 153 (7.9%) | 419 (8.4%) |
| Lives with kids youngest >=5 | 383 (17.3%) | 125 (14.9%) | 295 (15.2%) | 803 (16.1%) |
| **Education** |  |  |  |  |
| Certificate after 9 years | 380 (15.8%) | 81 (4.5%) | 276 (14.1%) | 737 (12.0%) |
| Certificate after 10 years | 569 (23.7%) | 149 (8.3%) | 341 (17.5%) | 1059 (17.2%) |
| Higher education | 1245 (51.8%) | 599 (33.2%) | 1296 (66.4%) | 3140 (51.0%) |
| None | 14 (0.6%) | 2 (0.1%) | 23 (1.2%) | 39 (0.6%) |
| Unclear | 197 (8.2%) | 973 (53.9%) | 15 (0.8%) | 1185 (19.2%) |
| **Occupation** |  |  |  |  |
| Employed | 1298 (59.2%) | 515 (61.7%) | 1103 (57.4%) | 2916 (58.9%) |
| Self employed | 138 (6.3%) | 75 (9.0%) | 137 (7.1%) | 350 (7.1%) |
| Unemployed | 6 (1.2%) | 12 (1.4%) | 36 (1.9%) | 74 (1.5%) |
| Retraining / Voluntary | 5 (0.2%) | 2 (0.2%) | 4 (0.2%) | 11 (0.2%) |
| Apprenticeship | 17 (0.8%) | 12 (1.4%) | 10 (0.5%) | 39 (0.8%) |
| Parental / other leave | 52 (2.4%) | 13 (1.6%) | 48 (2.5%) | 113 (2.3%) |
| Pupil / Retired | 656 (29.9%) | 206 (24.7%) | 584 (30.4%) | 1446 (29.2%) |
| Missing | 213 (8.9%) | 969 (53.7%) | 29 (1.5%) | 1211 (19.7%) |
| Changes due to COVID |  |  |  |  |
| No changes | 1.361 (59.4%) | 1.523 (52.4%) | 1.164 (57.0%) | 7.216 (58.8%) |
| Work from home | 642 (26.7%) | 276 (15.3%) | 554 (28.4%) | 1472 (23.9%) |
| Reduction work | 412 (17.1%) | 118 (6.5%) | 179 (9.2%) | 709 (11.5%) |
| Increase work | 217 (9.0%) | 104 (5.8%) | 169 (8.7%) | 490 (8.0%) |
| Short time work | 341 (30.2%) | 66 (14.7%) | 122 (14.9%) | 529 (22.1%) |
| Termination | 17 (1.5%) | 8 (1.8%) | 12 (1.5%) | 37 (1.5%) |
| New workplace | 31 (2.7%) | 15 (3.3%) | 42 (5.1%) | 88 (3.7%) |
| Other consequences | 189 (16.8%) | 60 (13.4%) | 119 (14.5%) | 368 (15.4%) |

**Table 3:**
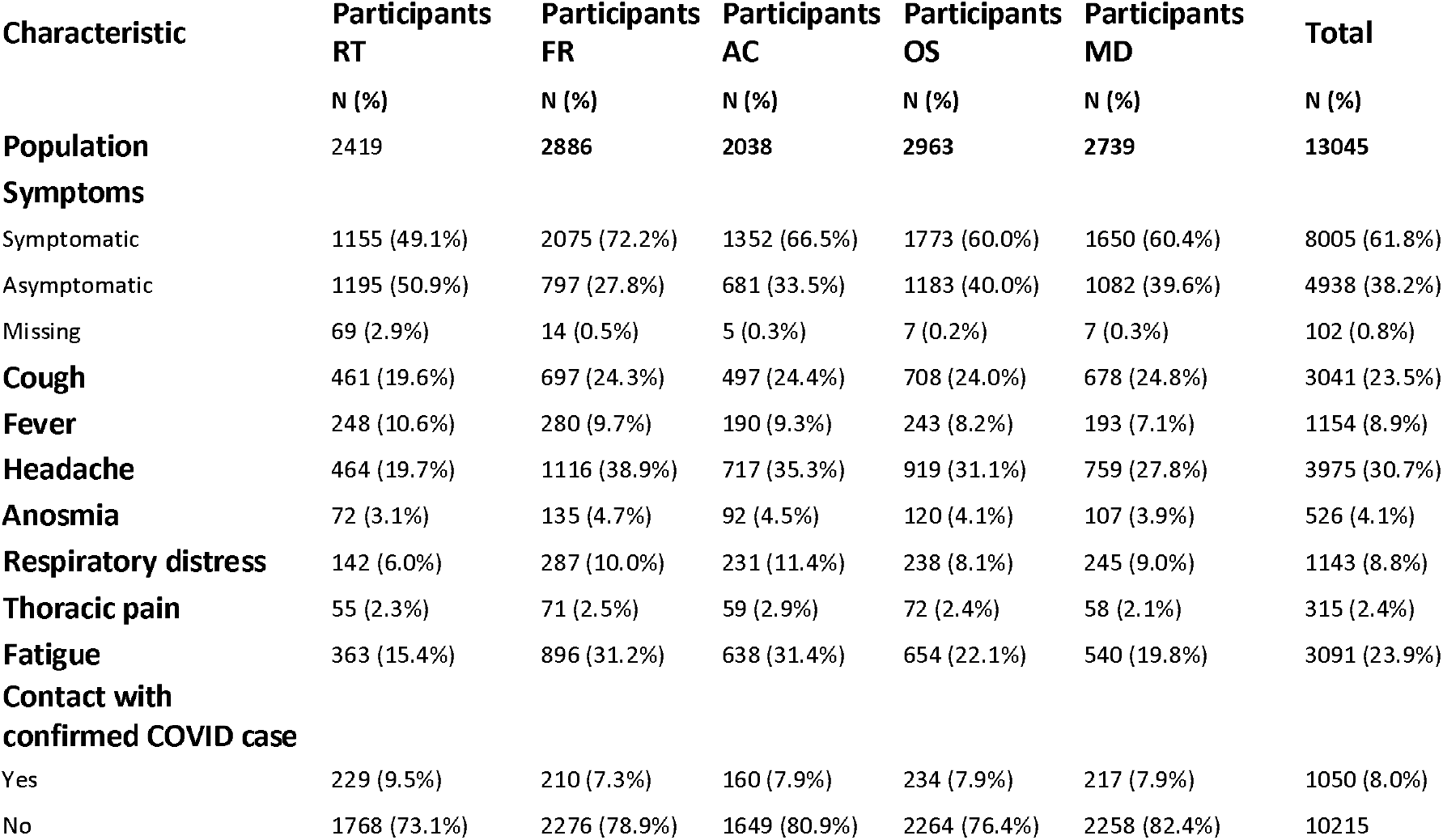
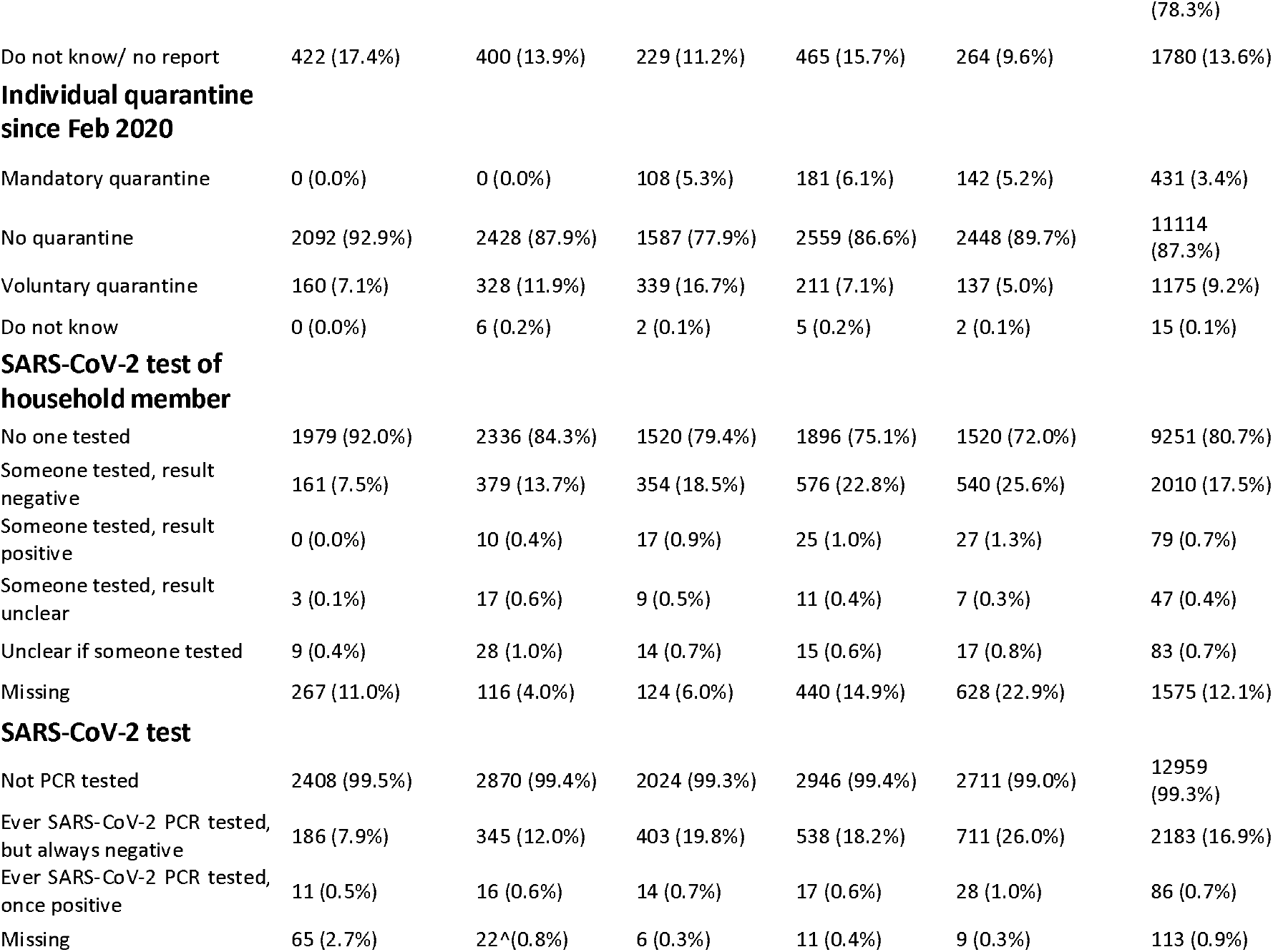
Overview of self-reported COVID-19 symptomatic and measures of exposure for five study population (Reutlingen (RT), Freiburg (FR), Aachen (AC), Osnabrück (OS) and Magdeburg (MD)) during phase 1, July-December 2020

**Table 4:** Overview of self-reported SARS-CoV-2 symptomatic and measures of exposure for three study population (Reutlingen (RT2), Freiburg (FR2), Aachen (AC2) during phase 2, October 2020 - February 2021

| Characteristic | Participants<br>RT 2 | Participants<br>FR 2 | Participants<br>AC 2 | Total |
| --- | --- | --- | --- | --- |
|  | N (%) | N (%) | N (%) | N (%) |
| **Population** | 2405 | 1804 | 1951 | 6160 |
| **Symptoms since Feb** |  |  |  |  |
| Symptomatic | 1282 (58.2%) | 585 (70.2%) | 1204 (61.9%) | 3071 (61.7%) |
| Asymptomatic | 922 (41.8%) | 248 (29.8%) | 740 (38.1%) | 1910 (38.3%) |
| Missing | 201 (8.4) | 971 (53.8) | 7 (0.4) | 1179 (19.1) |
| **Cough** | 482 (21.9%) | 208 (25.0%) | 482 (24.8%) | 1172 (23.5%) |
| **Fever** | 222 (10.1%) | 98 (11.8%) | 199 (10.2%) | 519 (10.4%) |
| **Headache** | 656 (29.8%) | 351 (42.1%) | 716 (36.8%) | 1723 (34.6%) |
| **Anosmia** | 90 (4.1%) | 40 (4.8%) | 112 (5.8%) | 242 (4.9%) |
| **Respiratory distress** | 163 (7.4%) | 81 (9.7%) | 219 (11.3%) | 463 (9.3%) |
| **Thoracic pain** | 43 (2.0%) | 25 (3.0%) | 70 (3.6%) | 138 (2.8%) |
| **Fatigue** | 26 (19.3%) | 270 (32.4%) | 529 (27.2%) | 1225 (24.6%) |
| **Contact with confirmed COVID case** |  |  |  |  |
| Yes | 193 (8.0%) | 110 (6.1%) | 328 (16.8%) | 631 (10.2%) |
| No | 1857 (77.2%) | 650 (36.0%) | 1526 (78.2%) | 4033 (65.5%) |
| Do not know/ no report | 355 (14.8%) | 1044 (57.9%) | 97 (5.0%) | 1496 (24.3%) |
| **Individual quarantine since Feb 2020** |  |  |  |  |
| Mandatory/ Voluntary quarantine | 357 (17.8%) | 192 (10.6%) | 417 (21.4%) | 966 (15.7%) |
| No quarantine | 1854 (83.7%) | 638 (76.8%) | 1531 (99.7%) | 4023 (87.8%) |
| Do not know | 3 (0.1%) | 1 (0.1%) | 5 (0.3%) | 9 (0.2%) |
| **SARS-CoV-2 test of household member** |  |  |  |  |
| No one tested | 1334 (73.0%) | 406 (60.6%) | 842 (53.3%) | 2582 (63.3%) |
| Someone tested, result negative | 453 (24.8%) | 233 (34.8%) | 654 (41.4%) | 1340 (32.9%) |
| Someone tested, result positive | 21 (1.1%) | 19 (2.8%) | 65 (4.1%) | 105 (2.6%) |
| Someone tested, result unclear | 11 (0.6%) | 4 (0.6%) | 2 (0.1%) | 17 (0.4%) |
| Unclear if someone tested | 9 (0.5%) | 8 (1.2%) | 17 (1.1%) | 34 (0.8%) |
| Missing | 577 (24.0) | 1134 (62.9) | 371 (19.0) | 2082 (33.8) |
| **SARS-CoV-2 test** |  |  |  |  |
| Not PCR tested | 1767 (80.1%) | 544 (65.8%) | 1034 (53.6%) | 3345 (67.4%) |
| Ever SARS-CoV-2 PCR tested, but always negative | 419 (19.0%) | 269 (32.5%) | 818 (42.4%) | 1506 (30.3%) |
| Ever SARS-CoV-2 PCR tested, once positive | 21 (1.0%) | 14 (1.7%) | 77 (4.0%) | 112 (2.3%) |
| Missing | 198 (8.2%) | 977 (54.2%) | 22 (1.1) | 1197 (19.4) |

### Known potential symptoms, exposure, quarantine and testing for SARS-CoV-2

Thirty-eight percent of participants in stage 1 and 2 had not experience any symptoms since February 2020 potentially related to SARS-CoV-2 at the time of data collection. Around nine percent had experienced fever (10.4% for stage 2), 23.5% cough (stage 1 and 2) and 23.9% fatigue (24.6% stage 2) respectively, and four percent anosmia (4.9% stage 2).

Eight percent (10.2% for stage 2) had known contact with a confirmed COVID-19 case and 12.6% (12% for stage 2) had been in voluntary or mandatory quarantine. Household members of 18.2% of the participants (35.5% stage 2) and 17.6% of participants (32.6% stage 2) had ever been tested for SARS-CoV-2 with a PCR test. Of these, 86 (0.7%) participants in stage 1 had ever tested positive and 112 (2.3%) participants in stage 2. The proportion of household members ever tested for SARS-CoV-2 by PCR increased from 8% in Reutlingen in July 2020 to 45% in Aachen in February 2021.

### Seroprevalence estimates in counties

Crude seroprevalence estimates for stage 1 were 2.5% (95% CI 1.9-3.2) in Reutlingen, 1.6% (95% CI 1.1-2.1) in Freiburg, 2.2% in Aachen (95% CI 1.6-3.0), 1.4% (95% CI 0.9-1.8) in Osnabrück and 2.3% (95% CI 1.8-3.0) in Magdeburg.

For stage 1 weighted seroprevalence estimates were 2.4% in Reutlingen (July 2020), 1.5% in Freiburg (August 2020), 2.3% in Aachen (September 2020), 1.4% in Osnabrück (October 2020) and 2.4% in Magdeburg (November-December 2020). In stage 2 we found an increase of weighted seroprevalence estimates over all study sites (RT2 in Oct/Nov 2020: 2.8%, FR2 in Nov/Dec 2020: 2.4% and AC2 Jan/Feb 2021: 5.5% excluding vaccinated participants vs. 7.1% including vaccinated participants (n=50)). In Reutlingen 2 in the age group 18-25 years we observed a decrease from the weighted seroprevalence 4.6% to 1.5%. The increase in weighted seroprevalence was highest among the > 79 age group. In Table 5 we report the unweighted seroprevalences.

**Table 5:** Unweighted and weighted SARS-CoV-2 seroprevalences for all study population (Reutlingen (RT), Freiburg (FR), Aachen (AC), Osnabrück (OS) and Magdeburg (MD)) July-February 2021

|  | Age groups | Crude for SARS-CoV-2 IgG ELISA | 95%CI | Population weighted for SARS-CoV-2 IgG ELISA | 95% CI |
| --- | --- | --- | --- | --- | --- |
| Reutlingen 1, July 2020 | all | 61 (2.5%) | 1.9 - 3.2 | 2.4% | 1.8 - 3.1 |
|  | 18-25 | 10 (4.2%) | 2.0 - 7.5 | 4.6% | 2.4 - 8.5 |
|  | 26-45 | 11 (1.8%) | 0.9 - 3.2 | 1.8% | 1.0 - 3.2 |
|  | 46-65 | 25 (2.5%) | 1.6 - 3.7 | 2.6% | 1.7 - 3.8 |
|  | 66-79 | 8 (2.3%) | 1.0 - 0.5 | 2.3% | 1.2 - 4.6 |
|  | >79 | 1 (0.9%) | 0.2 - 4.6 | 1.1% | 0.2 - 7.5 |
| Freiburg 1, August 2020 | all | 45 (1.6%) | 1.1 - 2.1 | 1.5% | 1.1 - 2.0 |
|  | 18-25 | 2 (0.8%) | 0.1 - 2.8 | 0.8% | 0.2 - 3.2 |
|  | 26-45 | 18 (1.8%) | 1.1 - 2.8 | 1.8% | 1.1 - 2.8 |
|  | 46-65 | 16 (1.6%) | 0.9 - 2.6 | 1.6% | 1.0 - 2.7 |
|  | 66-79 | 7 (1.6%) | 0.7 - 3.3 | 1.6% | 0.8 - 3.4 |
|  | >79 | 0 (0.0%) | 0 - 3.0 | 0.0% |  |
| **Aachen, September / October 2020** | all | 45 (2.2%) | 1.6 - 3.0 | 2.3% | 1.7-3.1 |
|  | 18-25 | 6 (3.0%) | 1.1 - 6.4 | 3.1% | 1.4 - 6.8 |
|  | 26-45 | 7 (1.2%) | 0.5 - 2.5 | 1.2% | 0.6 - 2.4 |
|  | 46-65 | 20 (2.5%) | 1.5 - 3.8 | 2.4% | 1.6 - 3.7 |
|  | 66-79 | 7 (2.0%) | 0.8 - 4.1 | 2.0% | 1.0 - 4.2 |
|  | >79 | 5 (5.3%) | 1.7 - 11.9 | 5.4% | 2.2 - 12.3 |
| **Osnabrück, October/November 2020** | all | 40 (1.4%) | 0.9 - 1.8 | 1.4% | 1.0 - 1.9 |
|  | 18-25 | 3 (1.3%) | 0.3 - 3.8 | 1.5% | 0.5 - 4.5 |
|  | 26-45 | 12 (1.5%) | 0.8 - 2.7 | 1.5% | 0.9 - 2.7 |
|  | 46-65 | 17 (1.3%) | 0.7 - 2.0 | 1.3% | 0.8 - 2.1 |
|  | 66-79 | 8 (1.8%) | 0.8 - 3.4 | 1.8% | 0.9 - 3.5 |
|  | >79 | 0 (0.0%) | 0 - 3.0 | 0.0% |  |
| **Magdeburg, November/December 2020** | all | 64 (2.3%) | 1.8 - 3.0 | 2.4% | 1.9 - 3.1 |
|  | 18-25 | 3 (2.1%) | 0.4 - 6.0 | 2.0% | 0.6 - 6.2 |
|  | 26-45 | 25 (3.1%) | 2.0 - 4.6 | 3.1% | 2.1 - 4.6 |
|  | 46-65 | 17 (1.8%) | 1.0 - 2.8 | 1.8% | 1.1 - 2.9 |
|  | 66-79 | 8 (1.4%) | 0.6 - 2.8 | 1.4% | 0.7 - 2.9 |
|  | >79 | 10 (4.7%) | 2.3 - 8.5 | 5.1% | 2.7 - 9.3 |
| **Reutlingen 2 October/ November 2020** | all | 61 (2.6%) | 2.0 - 3.3 | 2.8% | 2.1 - 3.7 |
|  | 18-25 | 4 (2.0%) | 0.1 - 5.0 | 1.5% | 0.6 - 4.0 |
|  | 26-45 | 12 (1.9%) | 1.0 - 3.3 | 2.0% | 1.1 - 3.6 |
|  | 46-65 | 34 (3.3%) | 2.2 - 4.6 | 3.4% | 2.4 - 4.7 |
|  | 66-79 | 8 (2.0%) | 0.9 - 3.8 | 2.0% | 1.0 - 4.0 |
|  | >79 | 3 (3.2%) | 0.7 - 9.0 | 5.7% | 1.9 - 16.1 |
| **Freiburg 2 November/ December 2020** | all | 40 (2.2%) | 1.6 - 3.0 | 2.4% | 1.8 - 3.4 |
|  | 18-25 | 4 (2.2%) | 0.6 - 5.9 | 2.2% | 0.8 - 5.9 |
|  | 26-45 | 17 (2.6%) | 0.9 - 3.1 | 2.8% | 1.7 - 4.4 |
|  | 46-65 | 12 (1.9%) | 0.6 - 4.2 | 2.0% | 1.2 - 3.6 |
|  | 66-79 | 5 (1.8%) | 0.4 - 1.3 | 1.8% | 0.8 - 4.3 |
|  | >79 | 2 (3.6%) | 0 - 3.0 | 4.4% | 1.1 - 15.8 |
| **Aachen 2 January / February 2021** | all | 137 (7.1%) | 5.9 - 8.3 | 7.1% | 6.0 - 8.4 |
|  | 18-25 | 19 (9.9%) | 5.9 - 14.7 | 9.6% | 6.2 - 14.6 |
|  | 26-45 | 32 (5.8%) | 4.0 - 8.1 | 5.4% | 3.8 - 7.6 |
|  | 46-65 | 61 (7.9%) | 6.1 - 10.0 | 8.0% | 6.2 - 10.1 |
|  | 66-79 | 18 (5.4%) | 3.2 - 8.4 | 5.3% | 3.4 - 8.3 |
|  | >79 | 7 (9.1%) | 3.7 - 17.8 | 8.8% | 4.2 - 17.6 |
| **Aachen 2, excluding vaccinated** | all | 102 (5.4%) | 4.4 - 6.5 | 5.5% | 4.5 - 6.8 |
|  | 18-25 | 12 (6.4%) | 3.3 - 10.8 | 6.5% | 3.7 - 11.2 |
|  | 26-45 | 20 (3.8%) | 2.3 - 5.7 | 3.5% | 2.2 - 5.4 |
|  | 46-65 | 47 (6.2%) | 4.5 - 8.0 | 6.4% | 4.9 - 8.5 |
|  | 66-79 | 16 (4.8%) | 2.8 - 7.7 | 4.8% | 3.0 - 7.6 |
|  | >79 | 7 (9.7%) | 4.0 - 19.0 | 9.4% | 4.4 - 18.7 |

### Seropositivity and confirmed SARS-CoV-2 infections

For stage 1 sixty-two percent (68/110) of those reporting ever having tested positive with a SARS-CoV-2 PCR had a positive antibody test in our sample. Nine percent (1/11) in Reutlingen, 25% (4/16) in Freiburg, in Aachen 35% (5/14), in Osnabrück 35% (6/17) and in Magdeburg 7% (2/28) were not detected by serological investigation. Of those with confirmed SARS-CoV-2 infections since February 2020 95% (65/68) suffered from symptoms compatible with COVID-19, while of those seropositive 74% (81/110) had symptoms (Supplement Table 1). For stage 2 80% (90/112) of those reporting ever having tested positive in a SARS-CoV-2 PCR had a positive antibody test.

### Surveillance Sensitivity

We express surveillance sensitivity as ratio of reported cases over infected cases as detected by our serological analysis. Ratios were 3.8 in Reutlingen, 2.2 in Freiburg, 4.9 in Aachen, 2.8 in Osnabrück and 5.1 in Magdeburg. For stage 2 the ratios were in 3.5 in Reutlingen 2, 2.2 in Freiburg 2, and 2.4 in Aachen 2. Surveillance detection ratio (SDR) across age groups were heterogeneous and had large CIs with lower detection ratios in the age group >79 (Figure 3B).

**Figure 3:**
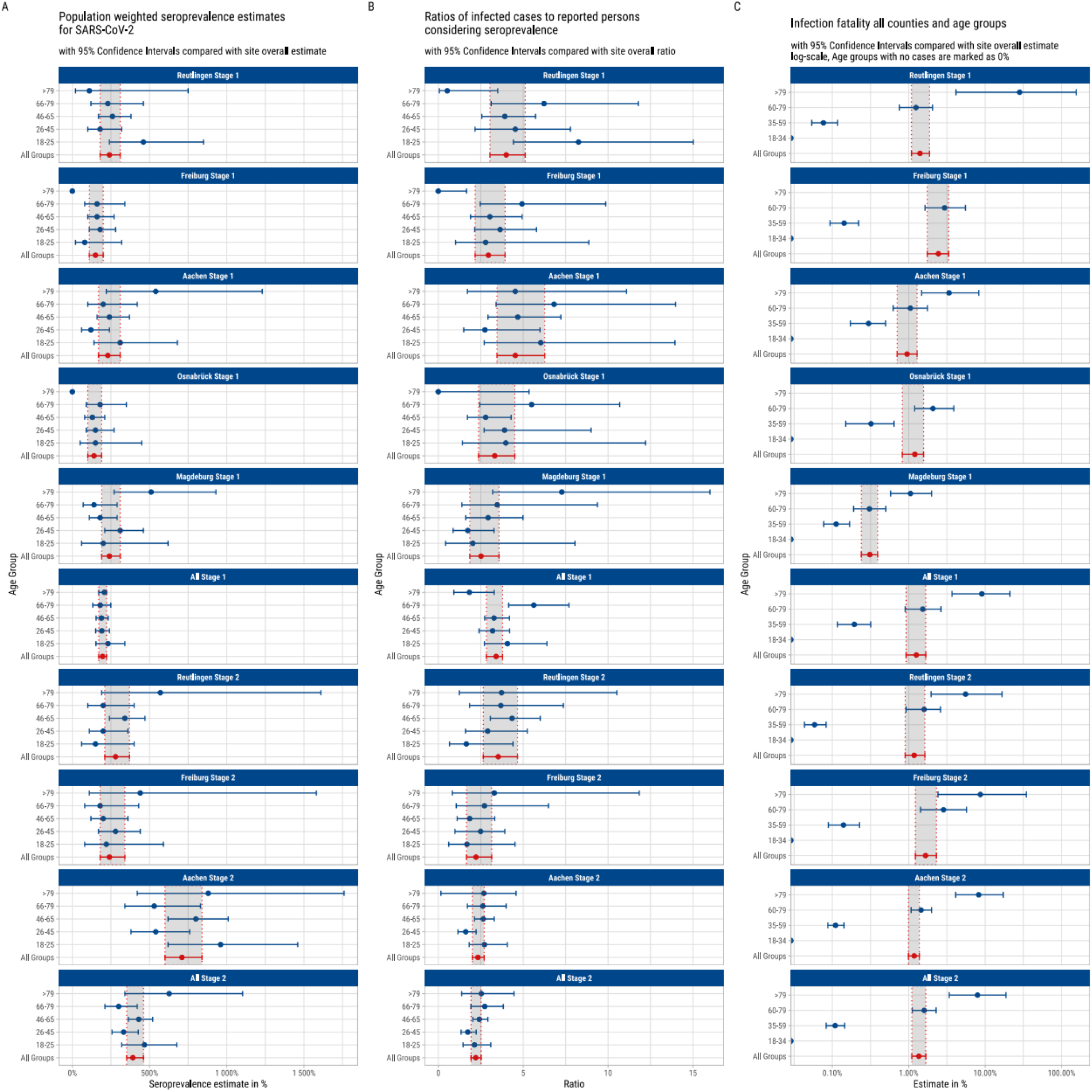
Stage 1 and 2 MuSPAD A: Population weighted seroprevalence estimates for SARS-CoV-2 B: Ratios of infected cases to reported persons considering seroprevalence C: Infection fatality rate counties and age group

### Infection fatality estimates

Infection fatality estimates were 1.4% in Reutlingen (95%CI 1.1-1.8%), 2.4% in Freiburg (95%CI 1.7-3.3%); 1% in Aachen (95%CI 0.7-1.3%), 1.2% in Osnabrück (95% CI 0.8-1.6%) and 0.3% in Magdeburg (95% CI 0.2-0.4%). Infection fatality estimates were highest in the oldest age group (>79) and decreased in younger age groups. In stage 2 the infection fatality estimates were 1.2% in Reutlingen2 (95%CI 0.9-1.6%); 1.7% in Freiburg 2 (95%CI 1.2-2.3%) and 1.2% in Aachen 2 (95%CI 1.0-1.4%) for more details see Figure 3 C and Table 6.

**Table 6:** Population weighted SARS-CoV-2 seroprevalence and estimated infection fatality ratio (IFR) by age groups and study sites in Germany, July 2020 to February 2021

|  | Seroprevalence | LCI95% | UCI95% | Inhabitants | Reported<br>COVID-19<br>deaths at<br>study start | Reported<br>COVID-19<br>cases 14<br>days before<br>study start | Infected based on<br>seroprevalence | Infected LCI95% | Infected UCI95% | IFR | IFR LCI95% | IFR UCI95% | crude CFR |
| --- | --- | --- | --- | --- | --- | --- | --- | --- | --- | --- | --- | --- | --- |
| **all RT** | **2.4%** | **1.8%** | **3.1%** | **237154** | **80** | **1492** | **5692** | **4269** | **7352** | **1.41%** | **1.09%** | **1.87%** | **5.36%** |
| 18-34 | 2.6% | 1.5% | 4.6% | 57802 | 0 | 262 | 1503 | 867 | 2659 | 0.00% | 0.00% | 0.00% | 0.00% |
| 35-59 | 2.6% | 1.7% | 3.7% | 100381 | 2 | 617 | 2610 | 1706 | 3714 | 0.08% | 0.05% | 0.12% | 0.32% |
| 60-79 | 2.3% | 1.4% | 3.8% | 59367 | 17 | 319 | 1365 | 831 | 2256 | 1.25% | 0.75% | 2.05% | 5.33% |
| >79 | 1.1% | 0.2% | 7.5% | 19604 | 61 | 294 | 216 | 39 | 1470 | 28.29% | 4.15% | 155.58% | 20.75% |
| **all FR** | **1.5%** | **1.1%** | **2.1%** | **412129** | **151** | **2091** | **6182** | **4533** | **8655** | **2.44%** | **1.74%** | **3.33%** | **7.22%** |
| 18-34 | 1.6% | 0.9% | 2.7% | 119900 | 0 | 489 | 1918 | 1079 | 3237 | 0.00% | 0.00% | 0.00% | 0.00% |
| 35-59 | 1.7% | 1.1% | 2.6% | 164717 | 4 | 859 | 2800 | 1812 | 4283 | 0.14% | 0.09% | 0.22% | 0.47% |
| 60-79 | 1.5% | 0.8% | 2.7% | 97329 | 43 | 418 | 1460 | 779 | 2628 | 2.95% | 1.64% | 5.52% | 10.29% |
| >79 | 0.0% | 0.0% | 0.0% | 30183 | 104 | 325 | 0 | 0 | 0 |  |  |  | 32.00% |
| **all AA** | **1.7%** | **3.1%** | **0.0%** | **470785** | **103** | **2193** | **10828** | **14594** | **8003** | **0.95%** | **0.71%** | **1.29%** | **4.70%** |
| 18-34 | 1.2% | 4.1% | 3.1% | 142227 | 0 | 586 | 3129 | 5831 | 1707 | 0.00% | 0.00% | 0.00% | 0.00% |
| 35-59 | 0.9% | 2.6% | 4.1% | 178758 | 8 | 906 | 2681 | 4648 | 1609 | 0.30% | 0.17% | 0.5% | 0.88% |
| 60-79 | 1.5% | 4.2% | 2.6% | 114037 | 30 | 427 | 2851 | 4790 | 1711 | 1.05% | 0.63% | 1.75% | 7.03% |
| >79 | 2.2% | 12.3% | 4.2% | 35763 | 65 | 274 | 1931 | 4399 | 787 | 3.37% | 1.48% | 8.26% | 23.72% |
| **all OS** | **1.3%** | **0.0%** | **1.9%** | **434567** | **68** | **2136** | **5649** | **4346** | **8257** | **1.20%** | **0.82%** | **1.56%** | **3.18%** |
| 18-34 | 1.5% | 0.7% | 0.0% | 116113 | 0 | 279 | 1742 | 1161 | 2206 | 0.00% | 0.00% | 0.00% | 0.00% |
| 35-59 | 1.4% | 0.8% | 2.6% | 177847 | 8 | 670 | 2490 | 1245 | 5335 | 0.32% | 0.15% | 0.64% | 1.19% |
| 60-79 | 1.5% | 0.0% | 0.0% | 105957 | 33 | 802 | 1589 | 848 | 2755 | 2.08% | 1.20% | 3.89% | 4.11% |
| >79 | 0.0% | 0.0% | 0.0% | 34650 | 40 | 190 | 0 | 0 | 0 |  |  |  | 21.05% |
| **all MD** | **2.2%** | **1.2%** | **3.8%** | **201596** | **15** | **948** | **4838** | **3830** | **6249** | **0.31%** | **0.24%** | **0.39%** | **1.58%** |
| 18-34 | 2.4% | 1.6% | 3.5% | 54425 | 0 | 905 | 1197 | 653 | 2068 | 0.00% | 0.00% | 0.00% | 0.00% |
| 35-59 | 1.8% | 1.1% | 2.9% | 74127 | 2 | 911 | 1779 | 1186 | 2594 | 0.11% | 0.08% | 0.17% | 0.22% |
| 60-79 | 5.1% | 2.7% | 9.3% | 54468 | 3 | 344 | 980 | 599 | 1580 | 0.31% | 0.19% | 0.50% | 0.87% |
| >79 | 0.0% | 0.0% | 0.0% | 18576 | 10 | 200 | 947 | 502 | 1728 | 1.06% | 0.58% | 1.99% | 5.00% |
| **all RT 2** | **2.9%** | **2.1%** | **3.8%** | **237154** | **81** | **1881** | **6877** | **4980** | **9012** | **1.18%** | **0.90%** | **1.63%** | **1.63%** |
| 18-34 | 2.0% | 1.0% | 4.0% | 57802 | 0 | 262 | 1156 | 578 | 2312 | 0.00% | 0.00% | 0.00% | 0.00% |
| 35-59 | 3.4% | 2.4% | 4.6% | 100381 | 2 | 617 | 3413 | 2409 | 4618 | 0.06% | 0.04% | 0.08% | 0.08% |
| 60-79 | 1.8% | 1.1% | 3.1% | 59367 | 17 | 319 | 1069 | 653 | 1840 | 1.59% | 0.92% | 2.60% | 2.60% |
| >79 | 5.7% | 1.9% | 16.1% | 19604 | 62 | 294 | 1117 | 372 | 3156 | 5.55% | 1.96% | 16.65% | 16.65% |
| **all FR 2** | **2.5%** | **1.8%** | **0.0%** | **412129** | **171** | **2091** | **10303** | **7418** | **14012** | **1.66%** | **1.22%** | **2.31%** | **2.31%** |
| 18-34 | 2.3% | 13.0% | 3.4% | 119900 | 0 | 489 | 2758 | 15587 | 4796 | 0.00% | 0.00% | 0.00% | 0.00% |
| 35-59 | 2.6% | 1.6% | 4.0% | 164717 | 6 | 859 | 4283 | 2635 | 6753 | 0.14% | 0.09% | 0.23% | 0.23% |
| 60-79 | 1.8% | 0.9% | 4.1% | 97329 | 50 | 418 | 1752 | 876 | 3504 | 2.85% | 1.43% | 5.71% | 5.71% |
| >79 | 4.4% | 1.1% | 3.6% | 30183 | 115 | 325 | 1328 | 332 | 4769 | 8.66% | 2.41% | 34.64% | 34.64% |
| **all AA 2** | **6.9%** | **5.9%** | **8.2%** | **470785** | **383** | **2.193** | **32484** | **27776** | **38604** | **1.18%** | **0.99%** | **1.38%** | **17.46%** |
| 18-34 | 6.0% | 4.2% | 8.6% | 142227 | 0 | 586 | 8534 | 5974 | 12232 | 0.00% | 0.00% | 0.00% | 0.00% |
| 35-59 | 7.6% | 5.9% | 9.6% | 178758 | 15 | 906 | 13586 | 10547 | 17161 | 0.11% | 0.09% | 0.14% | 1.66% |
| 60-79 | 6.6% | 4.8% | 8.9% | 114037 | 109 | 427 | 7526 | 5474 | 10149 | 1.45% | 1.07% | 1.99% | 25.53% |
| >79 | 8.8% | 4.2% | 17.6% | 35763 | 259 | 274 | 3147 | 1502 | 6294 | 8.23% | 4.11% | 17.24% | 94.53% |
| **all Aachen2 without vaccinated person** | **5.4%** | **4.4%** | **6.6%** | **470785** | **383** | **2.193** | **25422** | **20715** | **31072** | **1.51%** | **1.23%** | **1.85%** | **17.46%** |
| 18-34 | 3.9% | 2.4% | 6.2% | 142227 | 0 | 586 | 8534 | 5974 | 12232 | 0.00% | 0.00% | 0.00% | 0.00% |
| 35-59 | 5.5% | 4.1% | 7.4% | 178758 | 15 | 906 | 13586 | 10547 | 17161 | 0.11% | 0.09% | 0.14% | 1.66% |
| 60-79 | 5.9% | 4.2% | 8.2% | 114037 | 109 | 427 | 7526 | 5474 | 10149 | 1.45% | 1.07% | 1.99% | 25.53% |
| >79 | 9.4% | 4.4% | 18.7% | 35763 | 259 | 274 | 3147 | 1502 | 6294 | 8.23% | 4.11% | 17.24% | 94.53% |

### Association between measures of exposure to SARS-CoV-2 and risk of seropositivity

Overall the prevalence of SARS-CoV-2 IgG antibodies was 2% (254/13,045) for all sites between July and December 2020. 14.3% of those who had been ordered to mandatory quarantine were seropositive, in contrast to 1.2% of those who had not been in quarantine. This indicates 8.2 people have to quarantine to ensure that one infected person is in quarantine.

In a logistic regression participants reporting two of the typical COVID-19 symptoms (cough, anosmia, respiratory distress, fever and fatigue) during the last few months were associated with higher odds of being seropositive (aOR 3.6, 95%CI 3.2 - 4.3) (Tabel 7).

### Multivariable analysis of participant characteristics influencing risk of seropositivity

A logistic regression on population weighted seropositivity estimates accounting for clustering by study region showed evidence towards a higher risk of seropositive results in participants with lower education compared to higher education (aOR 1.8, 95%CI 1.2-2.7). There was lower risk of having a seropositive result in those who smoked daily (aOR 0.5, 95%CI 0.3-0.7). Age, rural or urban location, change and type of employment as well as further predisposing conditions did not change the odds of seropositivity in our analysis (Table 8).

**Table 7:**
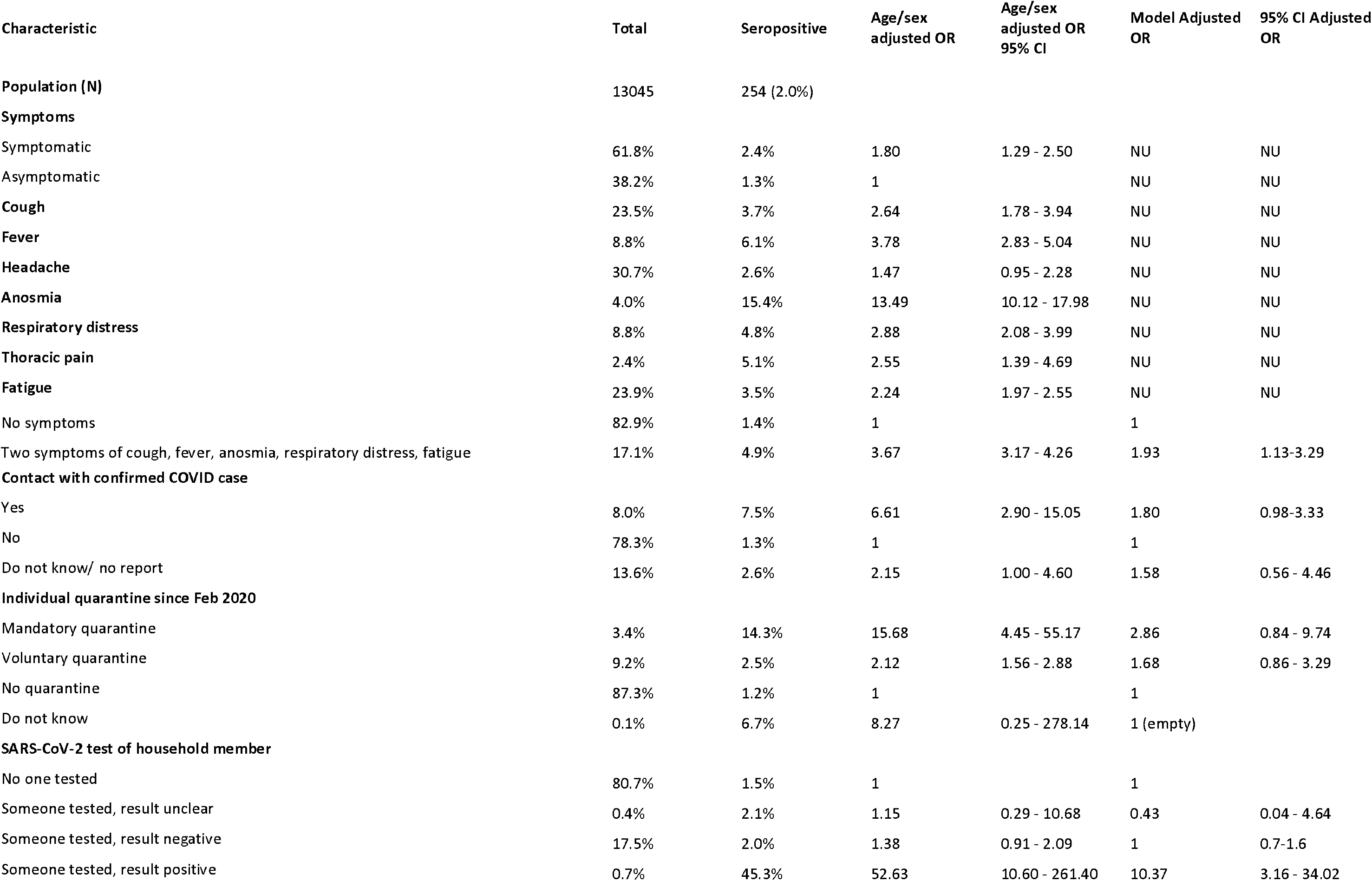
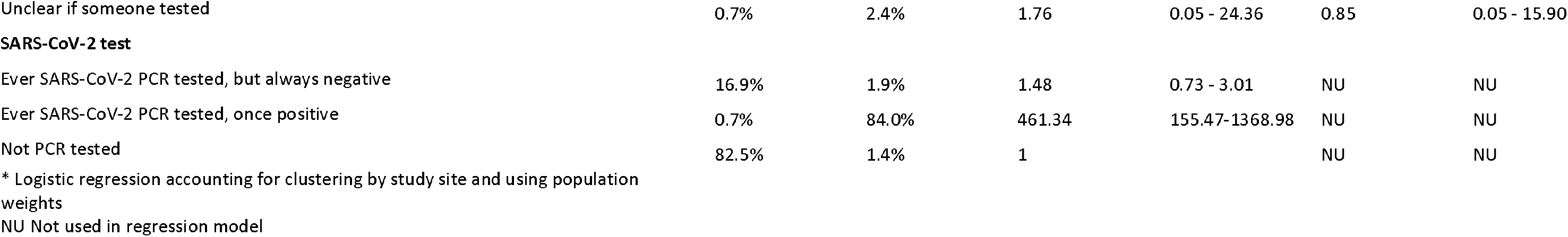
Logistic regression of measures of exposure to SARS-CoV-2 and their association with seropositivity for stage 1 of the five study centres, 2020

| Characteristic | Total | Seropositive | Age/sex<br>adjusted OR | Age/sex<br>adjusted OR<br>95% CI | Model Adjusted<br>OR | 95% CI Adjusted<br>OR |
| --- | --- | --- | --- | --- | --- | --- |
| **Population (N)** | 13045 | 254 (2.0%) |  |  |  |  |
| **Symptoms** |  |  |  |  |  |  |
| Symptomatic | 61.8% | 2.4% | 1.80 | 1.29 - 2.50 | NU | NU |
| Asymptomatic | 38.2% | 1.3% | 1 |  | NU | NU |
| **Cough** | 23.5% | 3.7% | 2.64 | 1.78 - 3.94 | NU | NU |
| **Fever** | 8.8% | 6.1% | 3.78 | 2.83 - 5.04 | NU | NU |
| **Headache** | 30.7% | 2.6% | 1.47 | 0.95 - 2.28 | NU | NU |
| **Anosmia** | 4.0% | 15.4% | 13.49 | 10.12 - 17.98 | NU | NU |
| **Respiratory distress** | 8.8% | 4.8% | 2.88 | 2.08 - 3.99 | NU | NU |
| **Thoracic pain** | 2.4% | 5.1% | 2.55 | 1.39 - 4.69 | NU | NU |
| **Fatigue** | 23.9% | 3.5% | 2.24 | 1.97 - 2.55 | NU | NU |
| No symptoms | 82.9% | 1.4% | 1 |  | 1 |  |
| Two symptoms of cough, fever, anosmia, respiratory distress, fatigue | 17.1% | 4.9% | 3.67 | 3.17 - 4.26 | 1.93 | 1.13-3.29 |
| **Contact with confirmed COVID case** |  |  |  |  |  |  |
| Yes | 8.0% | 7.5% | 6.61 | 2.90 - 15.05 | 1.80 | 0.98-3.33 |
| No | 78.3% | 1.3% | 1 |  | 1 |  |
| Do not know/ no report | 13.6% | 2.6% | 2.15 | 1.00 - 4.60 | 1.58 | 0.56 - 4.46 |
| **Individual quarantine since Feb 2020** |  |  |  |  |  |  |
| Mandatory quarantine | 3.4% | 14.3% | 15.68 | 4.45 - 55.17 | 2.86 | 0.84 - 9.74 |
| Voluntary quarantine | 9.2% | 2.5% | 2.12 | 1.56 - 2.88 | 1.68 | 0.86 - 3.29 |
| No quarantine | 87.3% | 1.2% | 1 |  | 1 |  |
| Do not know | 0.1% | 6.7% | 8.27 | 0.25 - 278.14 | 1 (empty) |  |
| **SARS-CoV-2 test of household member** |  |  |  |  |  |  |
| No one tested | 80.7% | 1.5% | 1 |  | 1 |  |
| Someone tested, result unclear | 0.4% | 2.1% | 1.15 | 0.29 - 10.68 | 0.43 | 0.04 - 4.64 |
| Someone tested, result negative | 17.5% | 2.0% | 1.38 | 0.91 - 2.09 | 1 | 0.7-1.6 |
| Someone tested, result positive | 0.7% | 45.3% | 52.63 | 10.60 - 261.40 | 10.37 | 3.16 - 34.02 |
| Unclear if someone tested | 0.7% | 2.4% | 1.76 | 0.05 - 24.36 | 0.85 | 0.05 - 15.90 |
| **SARS-CoV-2 test** |  |  |  |  |  |  |
| Ever SARS-CoV-2 PCR tested, but always negative | 16.9% | 1.9% | 1.48 | 0.73 - 3.01 | NU | NU |
| Ever SARS-CoV-2 PCR tested, once positive | 0.7% | 84.0% | 461.34 | 155.47-1368.98 | NU | NU |
| Not PCR tested | 82.5% | 1.4% | 1 |  | NU | NU |
| * Logistic regression accounting for clustering by study site and using population weights |  |  |  |  |  |  |
| NU Not used in regression model |  |  |  |  |  |  |

**Table 8:**
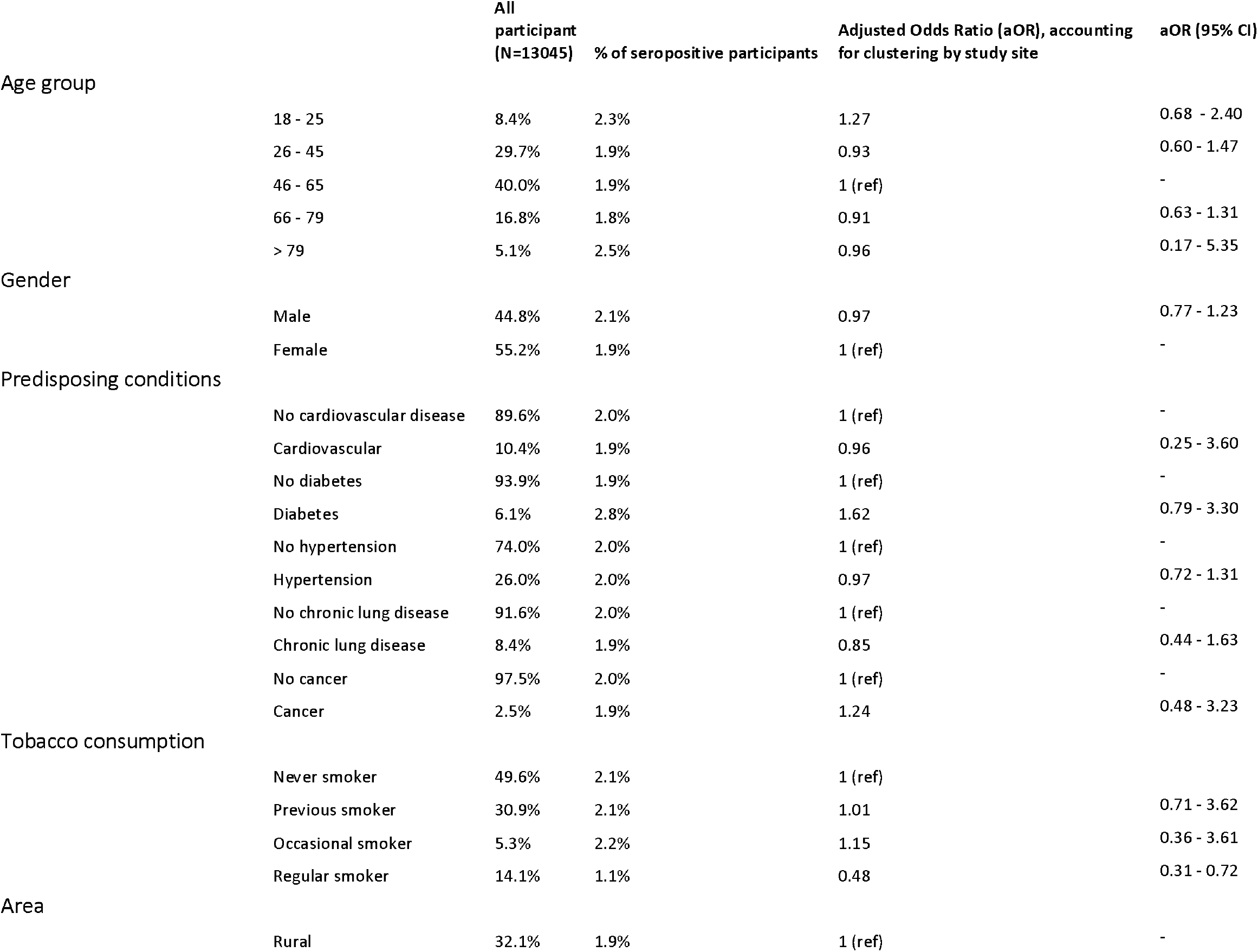
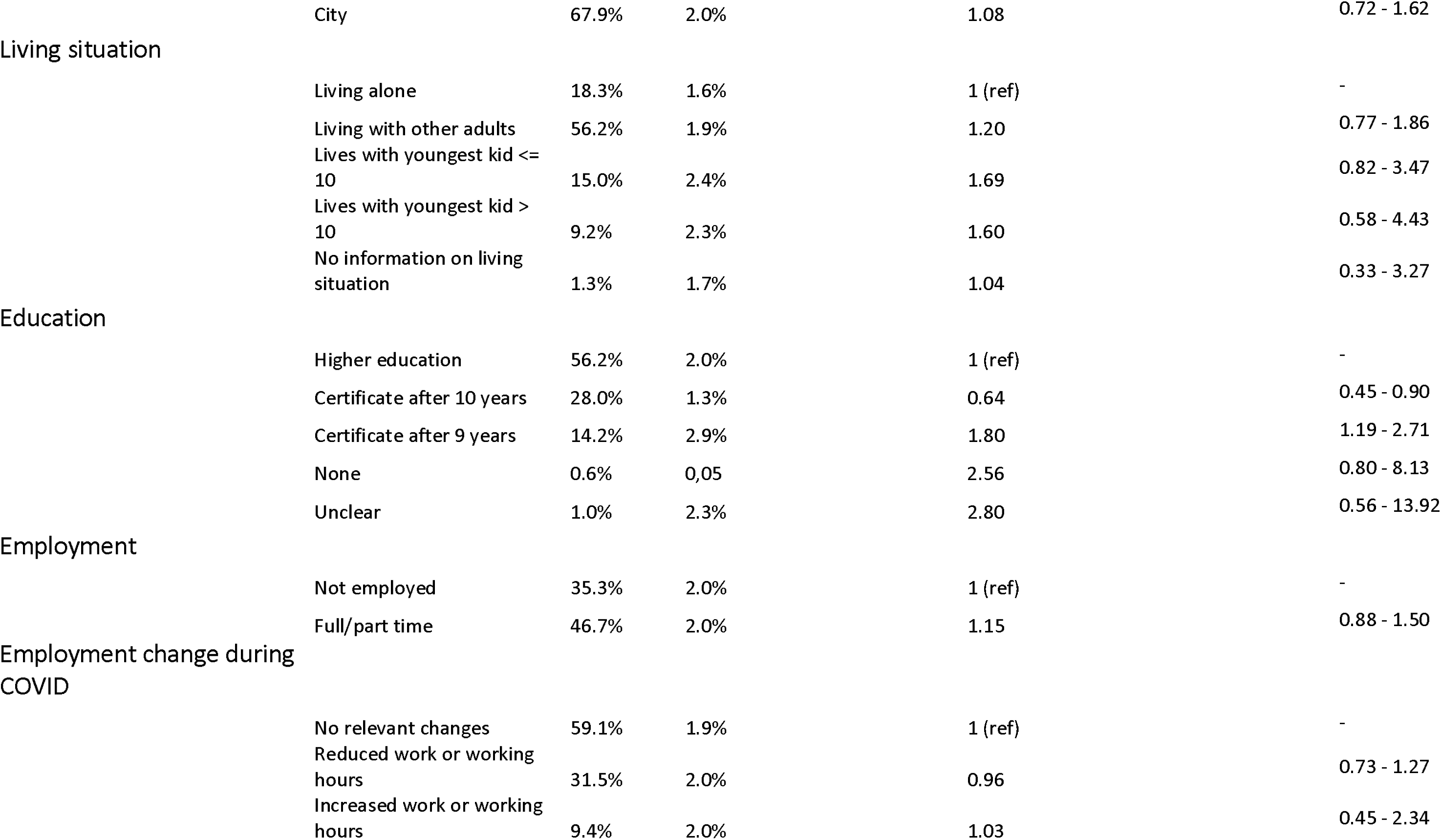
Multivariable analysis of MuSPAD participant characteristics influencing SARS-CoV-2 seropositivity for Reutlingen, Freiburg, Aachen, Osnabrück and Magdeburg stage 1, July-December 2020

## Discussion

Underreporting of cases and deaths during the COVID-19 pandemic has been a major problem of public health systems around the world^4^. We present first results from five counties in Germany of an ongoing population-based seroprevalence study. Weighted seroprevalence estimates ranged from 1.4% to 2.5% in different German regions between July and December 2020 and rose to 5.4% in February 2021. Detection ratios as a measure of surveillance infection sensitivity of those reported to authorities over those infected calculated from seroprevalence ranged from 2.5 to 4.5. Detection ratios were lower between December 2020 and February 2021 than during the first months of the pandemic. Infection fatality estimates ranged from 0.2 to 2.4%. The number needed to quarantine to ensure that one infected person was in quarantine was eight in the first wave.

Strengths of our seroprevalence study include its large sample size per region, the multilocal approach, as well as the random population-based sampling strategy. Limitations include the serial design in that it does not measure seroprevalence in all places at the same time. This means that compared to the measurements from July 2020 (three months after the peak of the epidemic) more antibody waning has taken place in those regions sampled later and up to major infection dynamics of the second wave in October/ November 2020. An additional limitation is since we did not include individuals younger than 18 years of age, our results can only be generalised for the adult population. All study materials were only provided in German and sufficient knowledge of the language was required to give written informed consent. Hence, this study cannot detect possible differences in the subgroup of the German population that does not fulfil these requirements.

Still, we believe that five main conclusions can be drawn from the estimates presented:

First, our estimates of both seroprevalence and our infection fatality estimates are in line with those few other available population-based reports and reports based on blood donor samples from Germany. These existing seroprevalence studies indicate low seroprevalences between 0.4 to 1.4% with an underreporting ratio of 2 to 8 and infection fatality estimates of 0.5 to 1.5% in studies up to November 2020^10 20 21^. Pooling all current population-based estimates for Germany yields a pooled seroprevalence of 1.3% (0.9 - 1.7%) with high heterogeneity (I^2^: 92%)^22^. Underreporting estimates over time from both existing studies and MuSPAD estimates show proportions of infections detected by notifications to be 20 - 40% (mirroring detection ratios of 2.5-5 in our study) from July to December 2020. For the second wave the proportion of infections detected by notifications increased to 40-50% (mirroring detection ratios 2-2.5 in our study) in most age groups and sites.

Second, seroprevalences between study regions were not highly variableconsiderably different in our study. This is in contrast to early hotpot studies mostly conducted in smaller German communities highly affected by the pandemic indicating large regional differences^20^. For the initial sampling periods in Freiburg, Reutlingen, Aachen, and Osnabrück, the majority of reported cases that accounts for the determined population seroprevalence was infected during the first wave. In contrast most of the seroprevalence found in Magdeburg in November 2020 is likely to already represent the beginning of the second pandemic wave. Reutlingen and Freiburg similarly show already rising seroprevalences during November and December 2020 and rapidly rising seroprevalence in February in Aachen reflecting the second wave dynamic.

Third, there are few determinants of seropositivity in our study. Some evidence suggests that education is a determinant of higher risk of seropositivity. Low education and related factors (SES, change in employment) have previously been shown to explain the individual potential to isolate at home or not^23^. As previous studies have reported^24^ seroprevalence among smokers was lower than among non-smokers. The explanation is unclear and may both be behaviour or physiology related. This finding has to be seen in light of the strong evidence of smoking being a risk factor for progression to more severe course of disease ^25^.

Fourth, age-specific estimates provided here, such as underreporting ratios and infections fatality estimates are heterogeneous across regions. Regional estimates are therefore crucial to provide a clear picture to stakeholders. Our age-specific infection fatality estimates are very much in line with previously reported estimates. The low overall infection fatality estimate in Magdeburg is largely explained by seroprevalence reflecting the early rising of cases during the second wave and our cut-off point for reported deaths being too early to show actual infection fatality. We included 15 deaths up to 15 November 2020, but 48 were notified up to 31^st^ December 2020. To our knowledge, our study is the first to assess regional and age specific differences in Surveillance Detection Ratio ranging from 25% to 50% between cities and overall being 2-fold higher among > 79 year olds (first wave) compared to the rest of the adult population, meaning that older people were more easily detected by notifications than younger people. We believe this to be of relevance for improving forecasting and modelling efforts and guiding risk assessment on targeted prevention measures. In how far this age specific difference detected in our study explains fast spillover of epidemic activity during the second wave in several European countries into older age groups with relatively higher epidemic activity in younger age groups should be evaluated in future modelling efforts.

Fifth, we show high contact tracing efficiency with a number needed to quarantine of 8 for quarantine mandated by officials. This is in line with estimates of secondary attack rates ranging from 7.2% in those not living in the same household to 13.0% of those living in the same household^26^. In comparison to the number needed to quarantine of population wide contact measures this is low – there is little evidence to interpret how this compares to other countries and their contact tracing programs. In our analysis participants having had contact with a confirmed COVID-19 case were six times more likely to be seropositive compared to those without. Several mathematical studies^27-29^ showed that efficient contact tracing and case isolation reduce the COVID-19 risk. Furthermore, participants who reported a household member with positive SAR-CoV-2 PCR test result were highly associated (aOR 52.6) with SARS-CoV-2 seropositivity. All this implies that control measurement of physical distancing and mandatory quarantine are effective tools to prevent the spread of disease.

We have presented results from a large ongoing seroprevalence study from several regions in Germany with already more than 19.000 participants indicating overall low seroprevalence from July to December 2020 and rising seroprevalence from December 2020 to February 2021 with comparably high infection fatality risks in particular in the old. Given the heterogeneity in underreporting estimates, we recommend that forecasting efforts use regional age-specific underreporting ratios if available to predict severe courses of disease as well as deaths more in detail. We also recommend that testing and tracing efforts be targeted in particular to those younger and middle-aged adult groups that seem to be prone to highest underreporting in our study.

### Individual contributions

Gérard Krause had the idea for the study. Berit Lange was responsible for the epidemiological study design, including sample size calculation and data analysis plan. Gérard Krause, Stefanie Castell and Yvonne Kemmling provided input into study design. Gérard Krause, Berit Lange, Monika Strengert and Stefanie Castell sought funding for this study. Monika Strengert and Tobias Kerrinnes coordinated laboratory procedures, support and equipment for the study. Daniela Gornyk, Berit Lange, Manuela Harries, Monika Strengert, Tobias Kerrinnes and Gérard Krause drafted all associated documents aimed for ethical approval. Yvonne Kemmling and Daniela Gornyk coordinated ethical and data protection concept approval of this study. Henrike Maaß, Manuela Harries and Daniela Gornyk reviewed literature. Stephan Glöckner and Julia Ortmann explored, transformed and cleaned the collected data. Daniela Gornyk, Manuela Harries, Pilar Hernandez and Monika Strengert coordinate the study. Manuela Harries, Daniela Gornyk, Monika Strengert and Jana Heise organized data collection. Daniela Gornyk, Manuela Harries, Stephan Glöckner and Julia Ortmann coordinated the data management. Berit Lange, Daniela Gornyk, Manuela Harries drafted the first version of this manuscript. Berit Lange, Daniela Gornyk and Manuela Harries performed the data analysis. Barbora Kessel carried out the supplemental adjustment to test performance. Stephan Glöckner was responsible for the data visualisation.

Kerstin Frank, Knut Gubbe and Torsten Tonn supported laboratory procedures and analyses.

Gottfried Roller, Michael Ziemons and Oliver Kappert provided local support and logistical planning on the ground and supported the interpretation of the local data.

All authors provided input into the first draft manuscript and all authors revised subsequent versions.

## Data Availability

The data used for this study can be made available anonymized to other academic researchers. Available data variables shared with the applicant will be: study site information, recruitment status, essay information, bio sample type, demographic information, self-administered diagnostic anamneses and seroprevalence test results. For more details please contact. Academic institutions can apply for the data via. The serohub is the seroprevalence virtual research environment that stores case based information of the MuSPAD study and other (inter-)national studies. The application process includes the review of the targeted research question and research method. After approval, a link to share the data will be send to the applicant. Shared data will be in regards to international data standards (for more details see https://www.covid19dataportal.org/support-data-sharing-covid19 in csv format.

https://serohub.net/de/

## Acknowledgements

Firstly, we sincerely thank counties and cities (Reutlingen, Freiburg/Breisgau Hochschwarzwald, Aachen, Osnabrück and Magdeburg) supporting and endorsing our study. We sincerely thank colleagues from the local public health departments for providing data in their districts. We are grateful to Anne Ulrike Marzian, Angelika Rath, Christina Suckel and Nicole Grupe for entering the data and answering the participant’s questions and concerns. We thank our colleagues in the Plauen and Osnabrück laboratories for the laboratory analyses. We thank Astrid Hans for administrative support. We thank Neha Warikoo for providing software assistance. We thank Kevin Grigorian and the Johanniter-Unfall-Hilfe, and Tim Balz and BOS112 for collaborating with us. We also thank Armgard Zindler and Marco Krischok and IPSOS for excellent technical appointment assistance. Our greatest thanks goes to all participating persons for donating their blood and time.

