## supplementary material for "SARS-CoV-2 seroprevalence in Germany - a population based sequential study in five regions"

### Supplementary information MuSPAD

#### Supplementary Introduction

This supplementary material has been provided by the authors to give readers additional information about their work.

#### Geographic locations of MuSPAD test centres

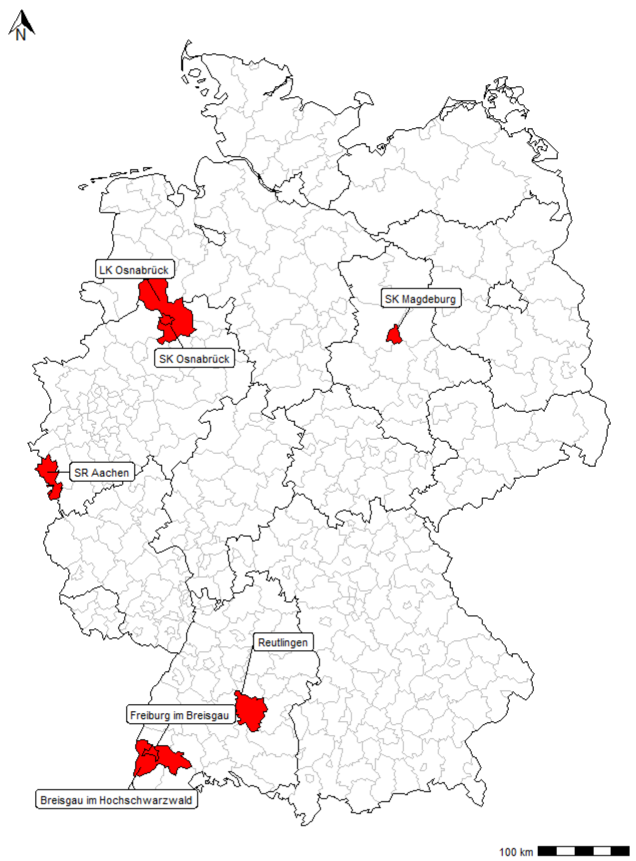

*Supplementary figure 1: Geographical distribution of the MuSPAD Sampling sites*

#### Supplementary Methods:

##### Data collection and management

###### Sampling

We invite all potential participants by postal mail including study information and instruction for booking an appointment through a booking service

###### Study procedures

The entire process follows Standardized Operating Procedures (SOPs). All those willing to participate have to give written informed consent. Field workers will register the participants in the study application "**P**rospective Monitoring and Management – **A**pp" (PIA[1]), and will take a blood sample (9 ml) and interview the participants using a standardised baseline questionnaire. To minimise the length of stay, we ask participants to answer a more detailed follow up questionnaire on e.g. health behaviour and living conditions from home.

#### Recruitment

##### Data collection

For each participant, questionnaires and blood sample are individually labelled with a barcode. The answers to the questionnaires are recorded in the secure web application PIA, which is already integrated in the “German National Cohort (NAKO)” and in the Cluster of Excellence “RESIST (Resolving Infection Susceptibility)” as electronic data capture system (ECD-System). The Helmholtz Centre for Infection Research (HZI) has developed PIA based on JavaScript and PostgreSQL, offering a hybrid mobile app (ionic framework) and a web-application for study participants’ surveys. Participants who prefer a paper-based version receive a postage-free return envelope containing a comprehensive questionnaire on the living situation, labelled with the individual barcode.
PIA offers specific web interfaces for all required professional roles (e.g. research team, participant management, study nurses). All questionnaire data as well as personal data of the participants can be recorded in PIA ensuring a strict separation of person-identifying data (idat) and medical/questionnaire data (mdat).
PIA also enables a longitudinal research with repeated surveys and sends out automatic reminders (push message or email) if questionnaires remain unanswered.

##### Interview/questionnaires

The field workers conduct an interview at the test centre using the baseline questionnaire. The interview takes approximately 10 minutes and incudes 14 questions on demographics, socioeconomic status, employment status, economic impact due to lockdown measures, comorbidities, smoking status, household, housing status, education level, flu like symptoms (self-reported), and other tests taken for SARS-CoV-2, and health status.

Follow-up questionnaires will allow us to track the health status of individuals and risk exposures. Details on pets and animals, hygiene related practices, prevention measures, medical history, hospitalisation, relevant medication and health seeking behaviour as well as work-life balance disruptions, psychological distress and level of concern over the pandemic are found in the follow up questionnaire.

##### Data management and data protection

We submitted a data protection concept to the national data protection authority (Federal Commissioner for Data Protection and Freedom of Information) and received approval.

The field workers commissioned and trained by the HZI enter the study data into the PIA database at the study site. Only field workers working on site and HZI researchers have access to the data, which are stored on HZI servers. Field workers do not have access to the laboratory data or the data from the follow-up questionnaire to ensure the separation of idat and mdat. The administration of the consent and the study data is carried out by HZI whereby a strict separation of person-identifying information as well as information from the survey and results of the blood samples is ensured. Pseudonymised laboratory data are stored in a separate laboratory database by the cooperation partners and the laboratory transmits the results of the analyses to the HZI at regular intervals. The laboratory records are merged with the PIA records using the participant’s identifiers. If participant changed their answers during the course of the interview, the last recorded answer in PIA is taken as the correct one. Laboratory data is checked for consistent reporting of the results as well as correct dates of analysis. Samples are compared with abnormalities documented by the laboratory. If samples were analyzed twice, the two results were manually compared and checked for discrepancies. In all of those cases, variation between quantitative results was only slight and did not influence the qualitative results, therefore the most recent result was considered correct. For the analysis, only participant with both a valid Euroimmun test result and at least partially filled in PIA questionnaires are considered. Since sex and birth dates are recorded at several points throughout the data collection process, these entries are checked for consistency and conflicts are set to NA. The age of the participant is calculated from the cleaned birth date information with a reference day being always the last date of the study period at the particular site. If only birth month and birth year of the participant is given, the 15th of the month is taken as the birth date, in case only the year is known,  June 15th is taken as the birth date for the participant. If the number of additional persons living in the same household is not directly given in the questionnaire, but persons (with or without details) are added in the questionnaire, their number is taken as a replacement for the missing information.

##### Sample preparation an dlab analysis

The blood is sampled in 9 ml EDTA-Monovettes, inverted carefully and stored at ambient temperature for 30 – 45 min prior to centrifugation with 2000x *g* for 10 minutes. The monovettes were stored at 4 - 8°C until transport to the central laboratory for selogical testing and aliquotation. Results are defined as positive, negative or borderline as classified by the test supplier. After the ELISA analysis, the remaining serum will be frozen at -80°C and stored at Hannover Unified Biobank HUB for further analysis.

### Supplementary Table(s) and Equation(s): Statistical Analysis Plan

#### Supplement Table 1: Seropositivity and confirmed cases

| age category | not detected | detected | total |
|---|---|---|---|
| 18-25 | 4 (50.0%) | 4 (50.0%) | 8 |
| 26-45 | 14 (38.9%) | 22 (61.1%) | 36 |
| 46-65 | 14 (34.2%) | 27 (65.9%) | 41 |
| 66-79 | 5 (38.5%) | 8 (61.5%) | 13 |
| >79 | 3 (37.5%) | 5 (62.5%) | 8 |
| total | 40 (37.7%) | 66 (62.3%) | 106 |

*Age category and PCR positives detected by serological assay, stage 1 MuSPAD participants*

#### Supplement Table 2: PCR positives stratified by sex

| gender | not detected | detected | total |
|---|---|---|---|
| female | 26 (41.3%) | 37 (58.7%) | 63 |
| male | 16 (34.0%) | 31 (66.0%) | 47 |
| total | 42 (38.2%) | 68 (61.8%) | 110 |

*Sex and PCR positives detected by serological assay, stage 1 MuSPAD participants*

#### Supplement Table 3: PCR positives stratified by site

| Self-reported PCR test (Once positive results) | not detected | Borderline | detected | Total |
|---|---|---|---|---|
| Reutlingen 2 | 4 (19.1%) | 0 (0%) | 31 (66.0%) | 27 |
| Freiburg 2 | 3 (21.4%) | 1 (7.1%) | 10 (71.4%) | 21 |
| Aachen 2 | 12 (15.5%) | 2 (2.6%) | 63 (81.8%) | 77 |

*Study site PCR positives detected by serological assay, stage 2 MuSPAD participants*

*:*

#### Supplement Table 4: Surveillance detection ratio (SDR) in all counties and overall

| | Seroprevalence (population weighted) | Seroprevalence LCI95% | Seroprevalence UCI95% | Reported cases 14 days before study start | Inhabitants | Infected | Infected LCI95% | Infected UCI95% | Detection ratio | Detection ratio 95%LCI | Detection ratio 95%LCI |
|---|---|---|---|---|---|---|---|---|---|---|---|
| Reutlingen 1 | | | | | | | | | | | |
| All | 2.4% | 1.8% | 3.1% | 1492 | 237154 | 5692 | 4269 | 7352 | 3.8 | 2.9 | 4.9 |
| 18-25 | 4.6% | 2.4% | 8.5% | 123 | 25965 | 623 | 623 | 2207 | 5.1 | 5.1 | 17.9 |
| 25-45 | 1.8% | 1,0% | 3.2% | 324 | 69966 | 3218 | 700 | 2239 | 9.9 | 2.2 | 6.9 |
| 46-65 | 2.6% | 1.7% | 3.8% | 572 | 84971 | 1529 | 1445 | 3229 | 2.7 | 2.5 | 5.6 |
| 66-79 | 2.3% | 1.2% | 4.6% | 179 | 36648 | 953 | 440 | 1686 | 5.3 | 2.5 | 9.4 |
| >79 | 1.1% | 0.2% | 7.5% | 294 | 19604 | 451 | 39 | 1470 | 1.5 | 0.1 | 5.0 |
| Freiburg 1 | | | | | | | | | | | |
| All | 2.4% | 1.8% | 3.4% | 4454 | 412129 | 9891 | 7418 | 14012 | 2.2 | 1.7 | 3.1 |
| 18-25 | 2.2% | 0.8% | 5.9% | 706 | 54091 | 1190 | 433 | 3191 | 1.7 | 0.6 | 4.5 |
| 25-45 | 2.8% | 1.7% | 4.4% | 1483 | 132072 | 3698 | 2245 | 5811 | 2.5 | 1.5 | 3.9 |
| 46-65 | 2,0% | 1.2% | 3.6% | 1458 | 134779 | 2696 | 1617 | 4852 | 1.8 | 1.1 | 3.3 |
| 66-79 | 1.8% | 0.8% | 4.3% | 404 | 61004 | 1098 | 488 | 2623 | 2.7 | 1.2 | 6.5 |
| >79 | 4.4% | 1.1% | 15.8% | 403 | 30183 | 1328 | 332 | 4769 | 3.3 | 0.8 | 11.8 |
| Aachen 1 | | | | | | | | | | | |
| All | 2.3% | 1.7% | 3.1% | 2193 | 470785 | 10828 | 8003 | 14594 | 4.9 | 3.6 | 6.7 |
| 18-25 | 3.1% | 1.4% | 6.8% | 270 | 70308 | 2180 | 984 | 4781 | 8.1 | 3.6 | 17.7 |
| 25-45 | 1.2% | 0.6% | 2.4% | 616 | 139203 | 1670 | 835 | 3341 | 2.7 | 1.4 | 5.4 |
| 46-65 | 2.4% | 1.6% | 3.7% | 795 | 153995 | 3696 | 2464 | 5698 | 4.6 | 3.1 | 7.2 |
| 66-79 | 2,0% | 1,0% | 4.2% | 238 | 71516 | 1430 | 715 | 3004 | 6.0 | 3.0 | 12.6 |
| >79 | 5.4% | 2.2% | 12.3% | 274 | 35763 | 1931 | 787 | 4399 | 7.0 | 2.9 | 16.1 |
| Osnabrück 1 | | | | | | | | | | | |
| All | 1.4% | 1,0% | 1.9% | 2136 | 434567 | 6084 | 4346 | 8257 | 2.8 | 2.0 | 3.9 |
| 18-25 | 1.5% | 0.5% | 4.5% | 279 | 54546 | 818 | 273 | 2455 | 2.9 | 1.0 | 8.8 |
| 25-45 | 1.5% | 0.9% | 2.7% | 670 | 128053 | 1921 | 1152 | 3457 | 2.9 | 1.7 | 5.2 |
| 46-65 | 1.3% | 0.8% | 2.1% | 802 | 152151 | 1978 | 1217 | 3195 | 2.5 | 1.5 | 4.0 |

| | | | | | | | | | | | |
|---|---|---|---|---|---|---|---|---|---|---|---|
| 66-79 | 1.8% | 0.9% | 3.5% | 190 | 65167 | 1173 | 587 | 2281 | 6.2 | 3.1 | 12.0 |
| >79 | 0,0% | | | 195 | 34650 | 0 | 0 | 0 | 0.0 | 0.0 | 0.0 |
| Magdeburg 1 | | | | | | | | | | | |
| All | 2.4% | 1.9% | 3.1% | 948 | 201596 | 4838 | 3830 | 6249 | 5.1 | 4.0 | 6.6 |
| 18-25 | 2,0% | 0.6% | 6.2% | 149 | 22952 | 459 | 138 | 1423 | 3.1 | 0.9 | 9.6 |
| 25-45 | 3.1% | 2.1% | 4.6% | 424 | 61890 | 1919 | 1300 | 2847 | 4.5 | 3.1 | 6.7 |
| 46-65 | 1.8% | 1.1% | 2.9% | 256 | 61402 | 1105 | 675 | 1781 | 4.3 | 2.6 | 7.0 |
| 66-79 | 1.4% | 0.7% | 2.9% | 57 | 36776 | 515 | 257 | 1067 | 9.0 | 4.5 | 18.7 |
| >79 | 5.1% | 2.7% | 9.3% | 62 | 18576 | 947 | 502 | 1728 | 15.3 | 8.1 | 27.9 |
| All | 2.0% | 1.7% | 2.2% | 8860 | 1756231 | 34422 | 30032 | 38988 | 3.9 | 3.4 | 4.4 |
| 18-25 | 2.3% | 1.5% | 3.4% | 1001 | 227862 | 5264 | 3486 | 7747 | 5.3 | 3.5 | 7.7 |
| 26-45 | 1.9% | 1.5% | 2.4% | 2612 | 531184 | 10146 | 8021 | 12748 | 3.9 | 3.1 | 4.9 |
| 46-65 | 1.9% | 1.5% | 2.3% | 3190 | 587298 | 11100 | 9044 | 13625 | 3.5 | 2.8 | 4.3 |
| 66-79 | 1.8% | 1.3% | 2.5% | 907 | 271111 | 4907 | 3579 | 6751 | 5.4 | 3.9 | 7.4 |
| >79 | 2.1% | 1.7% | 2.2% | 1150 | 138776 | 2873 | 2373 | 3081 | 2.5 | 2.1 | 2.7 |
| Reutlingen 2 | | | | | | | | | | | |
| All | 2.8% | 2.1% | 3.7% | 1881 | 237154 | 6640 | 4980 | 8775 | 3.5 | 2.6 | 4.7 |
| 18-25 | 1.5% | 0.6% | 4,0% | 236 | 25965 | 389 | 156 | 1039 | 1.7 | 0.7 | 4.4 |
| 26-45 | 2,0% | 1.1% | 3.6% | 481 | 69966 | 1399 | 770 | 2519 | 2.9 | 1.6 | 5.2 |
| 46-65 | 3.4% | 2.4% | 4.7% | 665 | 84971 | 2889 | 2039 | 3994 | 4.3 | 3.1 | 6.0 |
| 66-79 | 2,0% | 1,0% | 4,0% | 199 | 36648 | 733 | 366 | 1466 | 3.7 | 1.8 | 7.4 |
| >79 | 5.7% | 1.9% | 16.1% | 300 | 19604 | 1117 | 372 | 3156 | 3.7 | 1.2 | 10.5 |
| Freiburg 2 | | | | | | | | | | | |
| All | 2.4% | 1.8% | 3.4% | 4454 | 412129 | 9891 | 7418 | 14012 | 2.2 | 1.7 | 3.1 |
| 18-25 | 2.2% | 0.8% | 5.9% | 706 | 54091 | 1190 | 433 | 3191 | 1.7 | 0.6 | 4.5 |
| 26-45 | 2.8% | 1.7% | 4.4% | 1483 | 132072 | 3698 | 2245 | 5811 | 2.5 | 1.5 | 3.9 |
| 46-65 | 2,0% | 1.2% | 3.6% | 1458 | 134779 | 2696 | 1617 | 4852 | 1.8 | 1.1 | 3.3 |
| 66-79 | 1.8% | 0.8% | 4.3% | 404 | 61004 | 1098 | 488 | 2623 | 2.7 | 1.2 | 6.5 |
| >79 | 4.4% | 1.1% | 15.8% | 403 | 30183 | 1328 | 332 | 4769 | 3.3 | 0.8 | 11.8 |
| Aachen 2 | | | | | | | | | | | |

| | | | | | | | | | | | |
|---|---|---|---|---|---|---|---|---|---|---|---|
| All | 7.1% | 6,0% | 8.4% | 14118 | 470785 | 33426 | 28247 | 39546 | 2.4 | 2.0 | 2.8 |
| 18-25 | 9.6% | 6.2% | 14.6% | 2355 | 70308 | 6750 | 4359 | 10265 | 2.9 | 1.9 | 4.4 |
| 26-45 | 5.4% | 3.8% | 7.6% | 4562 | 139203 | 7517 | 5290 | 10579 | 1.6 | 1.2 | 2.3 |
| 46-65 | 8,0% | 6.2% | 10.1% | 4535 | 153995 | 12320 | 9548 | 15553 | 2.7 | 2.1 | 3.4 |
| 66-79 | 5.3% | 3.4% | 8.3% | 1253 | 71516 | 3790 | 2432 | 5936 | 3.0 | 1.9 | 4.7 |
| >79 | 8.8% | 4.4% | 17.6% | 1413 | 35763 | 3147 | 1574 | 6294 | 2.2 | 1.1 | 4.5 |
| All stage 2 | | | | | | | | | | | |
| all2 | 3.9% | 3.5% | 4.6% | 20453 | 1120068 | 43907 | 39426 | 51635 | 2.1 | 1.9 | 2.5 |
| 18-25 | 4.7% | 3.2% | 6.8% | 3297 | 150364 | 7022 | 4812 | 10180 | 2.1 | 1.5 | 3.1 |
| 26-45 | 3.3% | 2.6% | 4.3% | 6526 | 341241 | 11329 | 8770 | 14571 | 1.7 | 1.3 | 2.2 |
| 46-65 | 4.3% | 3.6% | 5.2% | 6658 | 373745 | 16071 | 13567 | 19435 | 2.4 | 2.0 | 2.9 |
| 66-79 | 3,0% | 2.1% | 4.2% | 1856 | 169168 | 5075 | 3569 | 7105 | 2.7 | 1.9 | 3.8 |
| >79 | 6.3% | 3.4% | 11.0% | 2116 | 85550 | 5364 | 2909 | 9436 | 2.5 | 1.4 | 4.5 |

#### Sensitivity analysis including test performance

To adjust for the unknown sensitivity and specificity of the Euroimmun test, we employed a Bayesian hierarchical model as outlined in [2] and modelled the number of positive test results in the different age and sex categories ($y_{agexsex}$) as coming from a binomial distribution with parameters $n_{agexsex}$and $p_{agexsex}$

$$y_{agexsex} \sim Binomial(n_{agexsex}, p_{agexsex})$$

where $n_{agexsex}$is the number of probands of the specific sex in the specific age category in our sample and $p_{agexsex}$denotes the apparent prevalence in the *age x sex* subgroup. We analysed each county separately.

The apparent prevalence is related to the "true" prevalence $\pi_{agexsex}$by accounting for the influence of sensitivity and specificity through

$$p_{agexsex} = \pi_{agexsex} se + (1 - \pi_{agexsex})(1 - sp)$$

where *se* and *sp* denote the sensitivity and specificity of the test in our study. For the logit-transformed age-and-sex specific prevalences we assumed a normal distribution with age specific mean and a common standard deviation

$$logit(\pi_{agexsex}) \sim N(\mu_{age}, \sigma)$$

where $logit(p) = log(p/(1-p))$. For the hyperparameters $\mu_{age}, \sigma$we assumed normal N(-1,2) and half-normal $N^+$ (0,0.5) priors. This choice of priors implies placing more prior belief in the lower end of the (0,1) interval regarding the age-and-sex-specific prevalences.

For sensitivity and specificity of the Euroimmun test, we utilized the recently published results of a validation study in[3] , which found a sensitivity of 88.3% based on 222 samples and specificity 99,2% based on 513 samples. We interpreted this as 196 true positive samples out of 222 and 509 true negative samples from 513. The numbers of true positive and true negative samples were modelled as coming from a binomial distribution involving the respective sample size and the unknown sensitivity and specificity, respectively, as parameters. We used uniform priors on the unknown test characteristics. We also examined the adjustment to the above sensitivity (196/222) in combination with the specificity reported recently by the producer of the test (1339/1344 = 0.996)[4], and to a sensitivity of 87% (127/146) and a specificity of 97.5% (547/561) as observed in[5].

To fit the described hierarchical model we used *rstan* package[6] in R 3.6.3[7].

We ran four chains with 20000 iterations each, discarding the first 10000 as the burn-in period (with adapt_delta set to 0.99). This resulted in 40 000 samples from the posterior distributions for the age-and-sex specific prevalences.

In order to estimate population-adjusted seroprevalences in different subpopulations (e.g. a specific age group), we determined the proportions of people in the subpopulation belonging to our pre-defined age x sex groups and used these proportions as weights when combining the draws from the posterior distribution of the sensitivity/specificity-adjusted age-and-sex-specific prevalences. The resulting posterior distribution of the population-adjusted seroprevalence in the subpopulation of interest was summarized in terms of posterior mean and 95% highest posterior density credible interval as calculated using the HDInterval package[7]. The crude rates are given in Table 3 in the main article. Given the low crude seroprevalences observed in our study, accurate information on the test characteristics and especially on the specificity, is crucial for the adjustment. Only in case of a high specificity, the adjusted estimates can be kept away from zero.

#### Supplement Table 5: Seropositivity and confirmed cases

| | Sens/Spec [%] | Age: 18-25 | Age: 26-45 | Age: 46-65 | Age: 66-79 | Age: >79 | All |
|---|---|---|---|---|---|---|---|
| Reutlingen1 | 87.0/97.5 | 3.1 (0.3 – 6.2) | 0.7 (0.0 – 1.7) | 1.0 (0.0 – 2.1) | 1.2 (0.0 – 2.9) | 1.3 (0.0 – 3.5) | 1.2 (0.5 – 2.0) |
| | 88.3/ 99.2 | 4.0 (1.2 – 7.3) | 1.2 (0.1 – 2.5) | 1.9 (0.5 – 3.2) | 1.9 (0.1 – 3.8) | 1.4 (0.0 – 3.8) | 1.9 (0.9 – 2.8) |
| | 88.3/ 99.6 | 4.5 (1.7 – 7.6) | 1.7 (0.5 – 3.0) | 2.4 (1.2 – 3.6) | 2.3 (0.6 - 4.3) | 1.5 (0.0 – 3.9) | 2.3 (1.5 – 3.2) |
| Freiburg 1 | 87.0/97.5 | 0.7 (0.0 – 1.8) | 0.8 (0.0 – 1.6) | 0.6 (0.0 – 1.4) | 0.9 (0.0 – 2.1) | 1.0 (0.0 – 2.7) | 0.7 (0.3 – 1.3) |
| | 88.3/ 99.2 | 0.8 (0.0 – 2.1) | 1.3 (0.3 – 2.4) | 1.2 (0.2 – 2.2) | 1.4 (0.1 – 2.7) | 0.9 (0.0 – 2.6) | 1.2 (0.5 – 1.8) |
| | 88.3/ 99.6 | 1.0 (0.0 – 2.3) | 1.6 (0.7 – 2.7) | 1.5 (0.5 – 2.5) | 1.6 (0.3 – 3.1) | 0.9 (0.0 – 2.6) | 1.4 (0.8 – 2.0) |
| Aachen 1 | 87.0/97.5 | 2.1 (0.0 – 4.6) | 0.5 (0.0 – 1.3) | 1.2 (0.0 – 2.4) | 1.1 (0.0 – 2.6) | 4.7 (0.1 – 9.8) | 1.4 (0.6 – 2.2) |
| | 88.3/99.2 | 2.8 (0.2 – 5.5) | 0.8 (0.0 – 1.7) | 2.0 (0.6 – 3.4) | 1.6 (0.1 – 3.3) | 5.5 (0.8 – 10.7) | 2.0 (1.0 – 2.9) |
| | 88.3/ 99.6 | 3.2 (0.7 – 6.0) | 1.1 (0.1 - 2.1) | 2.4 (1.1 – 3.7) | 2.0 (0.4 – 3.8) | 6.0 (1.3 – 11.2) | 2.3 (1.5 – 3.2) |
| Osnabrück 1 | 87.0/97.5 | 1.1 (0.0 – 2.7) | 0.7 (0.0 – 1.5) | 0.4 (0.0 – 1.0) | 1.0 (0.0 – 2.2) | 1.1 (0.0 – 3.0) | 0.7 (0.3 – 1.2) |
| | 88.3/99.2 | 1.3 (0.0 – 3.0) | 1.1 (0.1 – 2.2) | 0.8 (0.0 – 1.5) | 1.4 (0.1 – 2.8) | 1.1 (0.0 – 3.0) | 1.1 (0.4 – 1.7) |
| | 88.3/99.6 | 1.5 (0.0 – 3.3) | 1.4 (0.4 – 2.5) | 1.0 (0.3 – 1.8) | 1.7 (0.4 – 3.2) | 1.0 (0.0 – 2.9) | 1.3 (0.7 – 1.9) |
| Magdeburg 1 | 87.0/97.5 | 1.7 (0.0 – 4.2) | 1.8 (0.3 – 3.4) | 0.6 (0.0 – 1.4) | 0.6 (0.0 – 1.5) | 4.0 (0.6 – 7.6) | 1.4 (0.7 – 2.2) |
| | 88.3/ 99.2 | 2.1 (0.0 – 4.8) | 2.7 (1.2 – 4.3) | 1.2 (0.1 - 2.2) | 1.0 (0.0 – 2.0) | 4.8 (1.5 – 8.4) | 2.0 (1.1 – 2.9) |
| | 88.3/99.6 | 2.4 (0.1 – 5.3) | 3.1 (1.7 – 4.6) | 1.6 (0.5 – 2.6) | 1.3 (0.2 – 2.4) | 5.3 (2.1 – 8.8) | 2.4 (1.6 – 3.3) |

*Age-sex standardized seroprevalences [%] adjusted for sensitivity and specificity of the Euroimmun test (considering borderline results as negative) using sensitivity and specificity from different sources. Presented are posterior means and 95% highest posterior density credible intervals*

#### Supplement Table 6: SARS-CoV-2 associated symptoms, quarantine and contacts with seropositivity, interaction with age

| **Gender** | **OR** | **LCI** | **UCI** |
|---|---|---|---|
| female | ref | | |
| male | 1.3 | 0.8 | 1.5 |
| **2 of symptoms in age category** | | | |
| female 18 - 25 | 1.6 | 0.9 | 3.1 |
| female 26 - 45 | 1.0 | 0.6 | 1.6 |
| female 46 - 65 | ref. | | |
| female 66 - 79 | 1.5 | 0.9 | 2.6 |
| female >79 | 1.3 | 0.6 | 3.0 |
| male 18 - 25 | 1.5 | 0.6 | 3.8 |
| male 26 - 45 | 2.3 | 1.4 | 3.9 |
| male 46 - 65 | 3.5 | 2.2 | 5.5 |
| male 66 - 79 | 4.2 | 2.1 | 8.5 |
| male >79 | 4.6 | 1.3 | 16.8 |
| **quarantine** | | | |
| mandatory | 5.1 | 3.3 | 7.8 |
| voluntary | 1.6 | 1.0 | 2.5 |
| **no individual quarantine** | ref | | |
| do not know | 4.5 | 1.0 | 20.6 |
| contact | | | |
| no contact | ref | | |
| contact with confirmed case | 1.6 | 1.0 | 2.6 |
| do not know | 1.8 | 1.2 | 2.6 |
| **anyone in household PCR tested** | | | |
| **not tested** | **ref** | | |
| tested, result unclear | 1.3 | 0.2 | 7.9 |
| tested, result negative | 1.0 | 0.7 | 1.5 |
| tested, result positive | 8.5 | 4.4 | 16.2 |
| unclear | 1.0 | 0.6 | 1.7 |

*Euroimmune test results*

#### Supplement Table 7: Logistic regression on SARS-CoV-2 seropositivity

| | OR | LCI | UCI |
|---|---|---|---|
| **Gender** | | | |
| female | ref. | | |
| male | 1.02 | 0.63 | 1.65 |
| **2 of symptoms** | | | |
| no | ref. | | |
| yes | 2.36 | 1.88 | 3.0 |
| **age category** | | | |
| 18 - 25 | 0.96 | 0.41 | 2.53 |
| 26 - 45 | 0.88 | 0.65 | 1.19 |

| | | | |
|---|---|---|---|
| 46 - 65 | ref. | | |
| 66 - 79 | 1.06 | 0.75 | 1.50 |
| > 79 | 1.19 | 0.35 | 3.98 |
| **quarantine*contact** | | | |
| mandatory quarantine*no contact | 3.14 | 0.72 | 13.78 |
| mandatory quarantine*contact with COVID-19 case | 6.99 | 2.21 | 22.17 |
| mandatory quarantine*do not know | 15.28 | 5.92 | 39.39 |
| voluntary quarantine*no contact | 1.19 | 0.35 | 4.08 |
| voluntary quarantine*do not know | 2.95 | 1.57 | 5.56 |
| no individual quarantine*no contact | ref. | | |
| no individual quarantine*contact with COVID-19 case | 2.61 | 1.66 | 4.11 |
| no individual quarantine*do not know | 1.46 | 0.81 | 2.65 |
| do not know*no contact | empty | | |
| do not know*contact with COVID-19 case | ref. (empty) | | |
| do not know*do not know | 47.73 | 5.07 | 449.23 |
| **anyone in household PCR tested** | | | |
| not tested | ref. | | |
| tested, result unclear | 0.52 | 0.16 | 16.41 |
| tested, result negative | 1.24 | 0.63 | 2.46 |
| tested, result positive | 12.27 | 8.92 | 16.22 |
| unclear | 1.27 | 0.34 | 4.65 |

*Association with contacts, symptoms, quarantine and interaction between contact and quarantine*
